# Regression vs. Medical LLMs: A Comprehensive Study for CVD and Mortality Risk Prediction

**DOI:** 10.64898/2026.03.11.26347789

**Authors:** Samuel Desire Kom Sande, Maciej Skorski, Martin Theobald, Jochen Schneider, Winfried März

## Abstract

Cardiovascular diseases (CVDs) remain the foremost cause of global morbidity and mortality, driving an urgent need for robust predictive tools that enable early detection and preventive intervention. Traditional regression-based models—such as linear and logistic regression, regression trees and forests, and Support Vector Machines (SVMs)—have long underpinned CVD risk estimation but often assume linear relationships, homogeneous effects across populations, and a limited number of predictors. Recent advances in regression, such as bagging and boosting, as well as Generative Artificial Intelligence (GenAI) and Large Language Models (LLMs) are increasingly shifting this paradigm.

In this paper, we review key developments in the context of both classic regression techniques and recent GenAI approaches, and we put a particular focus on openly available Medical LLMs (MedLLMs) in combination with few-shot prompting and classification finetuning. Based on the LURIC cardiovascular health study, we investigate a broad variety of biomarkers and risk factors under two different cohorts of 3,316 CVD risk patients who underwent coronary angiography in Germany between 1997 and 2000. Our results demonstrate that large, pretrained MedLLMs (70B) achieve up to 82% AUROC for 1-year all-cause mortality (1YM) prediction with optimized few-shot prompting, thus performing competitively with recent regression techniques and state-of-the-art methods from the medical literature such as CoroPredict, SMART and SCORE2. Smaller models (8B) can be finetuned to match or even surpass their larger counterparts as well as commercial models like ClaudeSonnet-4.5 and ChatGPT-5.2. Among all evaluated approaches, the best-performing boosting-based regression technique (CatBoost) and commercial LLM (Gemini-3-Flash) both achieve an AUROC of up to 85%. Further model-calibration and -stratification analyses reveal a systematic mortality over-prediction (ECE: 0.05–0.10) of MedLLMs, while Platt scaling effectively reduces such miscalibrations by 60–90%.

## 1 Introduction

### Background & Motivation

Generative Artificial Intelligence (GenAI) has rapidly transformed various domains, including the Life Sciences and Healthcare, where its applications continue to expand in scope and sophistication. In particular, Large Language Models (LLMs) based on the Generative Pretrained Transformer (GPT) [43] architecture have demonstrated remarkable capabilities in understanding, generating, and reasoning with complex textual and structural information. These advances have sparked a growing interest in leveraging such models for clinical and biomedical tasks, including clinical diagnoses [50] and decision support [28], as well as patient risk stratification [31].

Cardiovascular diseases (CVDs) remain a leading cause of mortality worldwide [11]. Accurately assessing a patient’s mortality risk following a CVD diagnosis is therefore of crucial clinical importance, as it enables early risk stratification and facilitates targeted interventions for high-risk patients. Several computational methods have been developed for mortality risk prediction in the context of CVDs [17, 41, 42]. Machine Learning (ML) algorithms, including Support Vector Machines (SVMs) and boosting-enhanced regression techniques (such as LightGBM [22], XGBoost [8] and AutoML [6]), have demonstrated strong predictive capabilities for CVD and mortality risk estimation. In recent studies, Artificial Neural Networks (ANNs) have been employed to predict 1-year mortality of patients following myocardial infarction (MI) [30] by training a 10-layer ANN on 21 carefully curated biomarkers derived from Electronic Health Records (EHRs). Over the past years, attention has increasingly turned toward LLMs and their potential applications in biomedical prediction tasks, including CVD [44, 49] and more general mortality risk estimation [15, 17], and even for predicting entire health trajectories with the goal of creating patients’ digital twins [29].

LLMs are generally based on the Transformer architecture [43], which employs a self-attention mechanism to process sequential input tokens and then calculates series of conditional probabilities to predict the most likely output tokens. Depending on which part of the Transformer architecture they utilize, LLMs can be broadly categorized into three families.

- **Encoder-based LLMs** leverage the encoder component of the Transformer architecture and are primarily applied to Natural Language Processing (NLP) tasks that involve understanding or representation learning, such as clustering and classification (e.g., for sentiment analysis). In the medical domain, encoder-based models—such as BERT (Bidirectional Encoder Representations from Transformers) [10]—have demonstrated strong potential. For example, multimodal finetuning of BERT has been successfully applied for predicting COVID-19 outcomes [16, 33]. However, encoder-based models are primarily discriminative and not inherently generative [21], limiting their flexibility for tasks involving reasoning, synthesis, or explanation generation— capabilities increasingly sought in clinical AI applications.
- **Decoder-based LLMs**, like LLaMA [40] and Qwen [2, 48], in contrast, utilize the decoder component of the Transformer architecture and are typically employed for generative tasks, such as dialogue systems and question answering. In clinical settings, decoder-based models are particularly suitable for explanatory or summarization tasks, as they are required for clinical diagnoses and decision making [1, 30]. Moreover, [4] demonstrated that decoder-based LLMs can even outperform their encoder-based counterparts in the quality of their text embeddings, which in turn makes them attractive also for finetuning and classification tasks. In our experiments (Section 4), we show that decoder-based models can indeed be turned into very effective risk predictors for CVD and mortality prediction.
- **Encoder-Decoder-based LLMs**, like DeepSeek [9], in contrast utilize both the encoder and decoder components of the Transformer architecture for language-generation tasks.

### Limitations of Current Approaches

Recent studies [15, 44, 49] mostly focus on applying commercial LLMs, such as ChatGPT, Claude or Gemini, for CVD prediction via specific prompting strategies. Typically, these studies employ EHR data in zero-shot settings, where models are directly applied without additional domain-specific finetuning. In [38], openly available LLMs (Qwen-2 & LlaMA-3), were tested for 1-year all-cause mortality prediction in MI patients, but their performance still significantly lagged behind that of the ANN proposed by [30]. Recent advances [1] in finetuning Medical LLMs^1^ (MedLLMs) demonstrated their potential to support clinical decision-making, showing that domain adaptation enhances both accuracy and reliability. Building on these results, [5] introduced the “Clinical Prediction with Large Language Models” (CPLLM) framework, which combines prompting and supervised finetuning (SFT) of pretrained MedLLMs to predict disease onset and hospital readmission using EHR data. These findings establish a strong precedent for leveraging LLMs as predictive and decision-support tools in Healthcare.

The usage of LLMs in clinical applications however relies strongly on the availability of textual EHRs or discharge summaries as their primary source for training and evaluation. While discharge notes provide rich clinical context, they are not consistently available across Healthcare domains and often remain expensive and time-consuming to produce, as they require extensive clinician input. This reliance poses a significant barrier to the scalability and generalizability of LLMs for medical applications.

### Contributions & Outline

Motivated by these limitations, our research explores an alternative and more cost-effective source of clinical information—routinely collected test data and comorbidity profiles. Such data are widely available across Healthcare settings and provide objective, structured indicators of a patient’s health status. We hypothesize that these routinely measured biomarkers can serve as a strong basis for predicting mortality risk in CVD patients. We therefore summarize our contributions as follows.

- Based on the LURIC cardiovascular health study (Section 3.1), we propose a wide range of **cost-effective biomarkers and comorbidities** (Section 3.2) as input both for classic ML algorithms (Section 3.3) and recent MedLLMs for CVD mortality risk predictions, over using expensive discharge notes.
- We propose a unified way to translate tabular patient records into **generic zero- and few-shot prompting schemes** (Section 3.4) which are suitable for CVD and mortality risk estimation under all—both commercial and openly available—LLMs we considered in the context of this study.
- We investigate **supervised finetuning** (SFT) techniques to further specialize pretrained MedLLMs for CVD and mortality risk estimation (Section 3.6).
- We provide the—to our knowledge—so far **most comprehensive study** of recent regression techniques, including boosting extensions of ML algorithms as well as Tabular Foundation Models, against both commercial and openly available LLMs (Section 4.5).
- Further extensions of our study include **model calibration and stratification** to reduce mortality over-estimations which frequently occur in these settings (Section 4.6).

## 2 Related Work

### 2.1 Encoder-Based LLMs in Medicine

With the advent of Transformer-based architectures [43], encoder-based LLMs such as BERT (Bidirectional Encoder Representations from Transformers) have become foundational tools for natural language understanding (NLU). Their applications span sentiment analysis [3], translation [51], summarization [27] and, increasingly, clinical tasks. For instance, multimodal finetuning of BERT has been used for COVID-19 outcome prediction and clinical text classification [16, 33]. These models are designed to encode input sequences into semantically meaningful vector representations, enabling effective learning across structured and unstructured health-care data. However, encoder-based models are primarily discriminative and not inherently generative [21], limiting their flexibility for tasks involving reasoning, synthesis, or explanation generation—capabilities increasingly sought in clinical AI systems.

### 2.2 Decoder-Based LLMs for Clinical Prediction

The recent emergence and success of decoder-based generative LLMs—including GPT [43], LLaMA [40], and Qwen[2, 48] —have expanded the scope of AI applications to include open-ended reasoning [13], question answering [36], and data-driven hypothesis generation [32]. Traditionally viewed as generators rather than classifiers, these models are now being evaluated for language understanding tasks previously dominated by encoder architectures. For example, [4] demonstrated that decoder-based LLMs can outperform their encoder-based counterparts in the quality of their text embeddings and other semantic tasks. In the medical domain, initial attempts to apply decoder-based LLMs to mortality prediction have yielded mixed results. In [38], open-source LLMs (LLaMA & Qwen) were tested for 1-year all-cause mortality prediction in MI patients, but their performance lagged behind that of the ANN proposed by [30]. Several factors contributed to this gap:

- The models were evaluated only in zero-shot settings, without domain adaptation.
- They were prompted on clinician-written discharge summaries rather than structured biomarker data.
- Being general-purpose models, they lacked comprehensive exposure to medical domain knowledge.

These limitations underscore the need for domain-specific fine-tuning and structured-data integration when deploying LLMs for clinical predictions.

### 2.3 Finetuning LLMs for Clinical Decision Support

Recent advances indicate that finetuning and prompt optimization can substantially improve LLM performance on specialized medical tasks. For example, [1] demonstrated the potential of finetuned LLMs to support clinical decision-making, showing that domain adaptation enhances both accuracy and reliability. Building on this, [5] introduced the “Clinical Prediction with Large Language Models” (CPLLM) framework, which combines prompting and supervised finetuning (SFT) of pre-trained LLMs to predict disease onset and hospital readmission using EHR data. CPLLM achieved state-of-the-art (SOTA) results across benchmarks, measured by Precision-Recall AUC (PR-AUC) and Receiver-Operating-Characteristic AUC (ROC-AUC) metrics. These findings establish a strong precedent for leveraging LLMs as predictive and decision-support tools in healthcare.

### 2.4 Research Gap & Motivation

Despite these promising developments, several critical gaps remain unaddressed:

- Most LLM-based mortality prediction studies rely on textual discharge summaries, which are costly and inconsistently available across healthcare systems.
- The use of routine, structured biomarkers as an alternative data source for LLMs has not yet been systematically investigated.
- The comparative effectiveness of decoder-based vs. encoder-based architectures for tabular biomedical prediction tasks remains largely unexplored.
- The integration of explainability mechanisms within Medical LLMs–crucial for clinician trust and transparency—has so far been explored only to a limited extent.

To bridge these gaps, our study investigates the potential of decoder-based Medical LLMs (MedLLMs) for 1-year all-cause mortality prediction in CVD patients using routinely measured biomarkers. We further explore few-shot prompting and supervised fine-tuning, to evaluate the performance, and clinical viability of these models relative to traditional ML and tabular foundation model baselines.

## 3 Methodology

Our methodology integrates classical Machine Learning (ML) algorithms, Tabular Foundation Models (TFMs), and Large Language Models (LLMs), enabling a comprehensive assessment of both traditional and emerging paradigms in clinical risk prediction.

The workflow consists of four major components as shown in Figure 1. First, we preprocess and structure the clinical biomarkers extracted from electronic health records (EHRs), ensuring consistent representations across models. Second, we train baseline ML methods and state-of-the-art TFMs to establish their reference performance on the given prediction task. Third, we design LLM-based prompting pipelines that translate biomarker sets into textual prompts suitable for in-context learning and few-shot prompting. Finally, we introduce an LLM classification finetuning pipeline to further extend this range of approaches.

**Figure 1.**
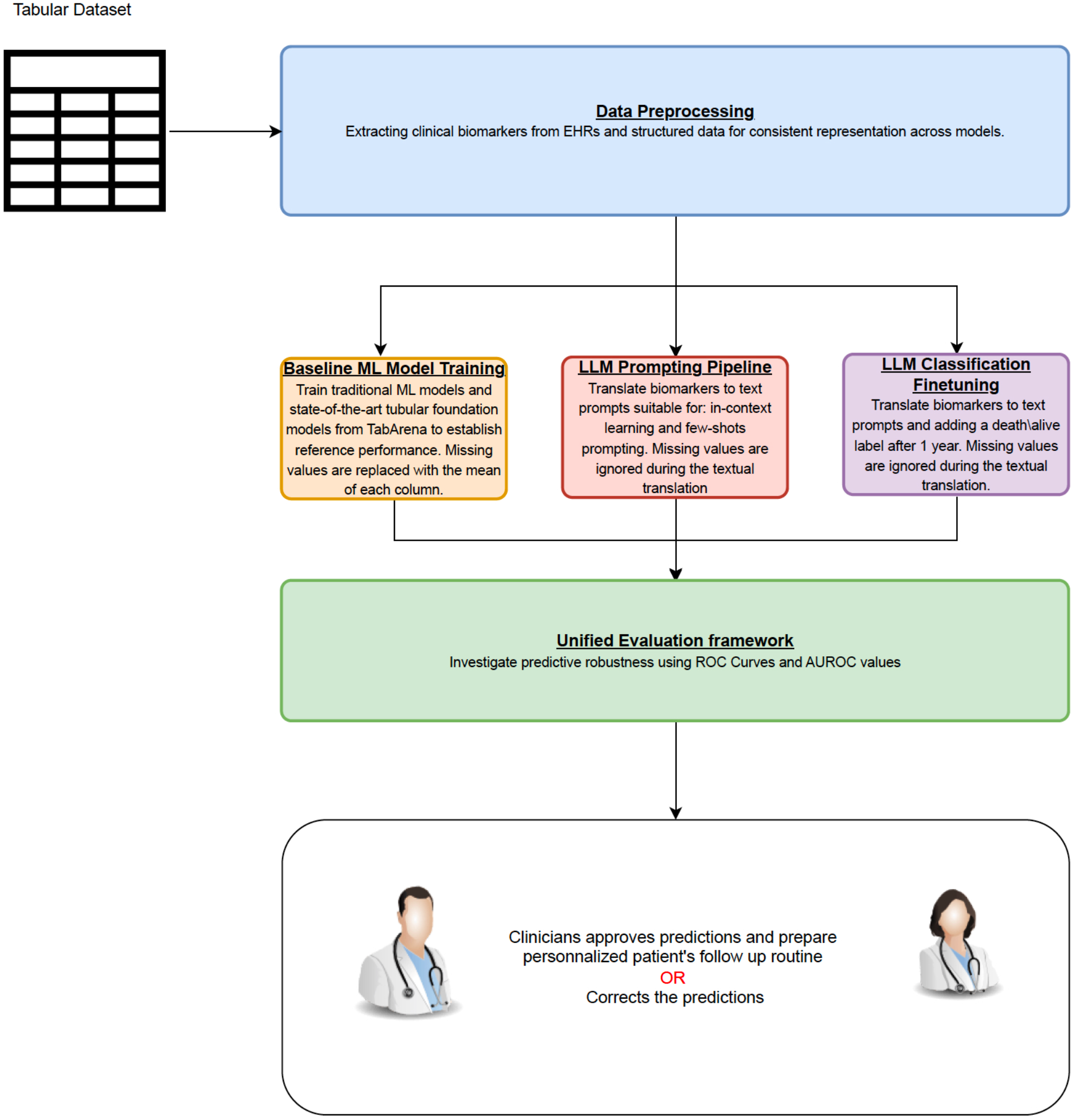
One-year all-cause mortality prediction workflow.

Together, these methodological components provide a unified evaluation framework that allows us to investigate the predictive capabilities, limitations, and practical value of LLMs compared with established ML approaches in clinical mortality prediction. A clinician acts as a final module who approves the predictions and establishes a targeted followup plan for the patient, or corrects the predictions. The following subsections describe each component in detail.

### 3.1 Study Design & Cohorts

We focus on the Ludwigshafen Risk and Cardiovascular Health (LURIC) study [45], which is a large, ongoing prospective investigation aimed at improving the prevention and management of CVD. A total of 3,316 patients were enrolled between 1997 and 2000, making LURIC one of the most comprehensive and best-characterized cardiovascular cohorts available. All participants were treated for CVD at the Ludwigshafen Hospital and underwent coronary angiography as part of their clinical evaluation.

In addition to standardized clinical assessments, participants also completed extensive questionnaires detailing dietary habits, lifestyle factors, and their medical history. The cohort was followed for an average of approximately 10 years, enabling robust evaluation of long-term health outcomes, including cardiovascular and allcause mortality. The richness of the phenotypic data, combined with the long follow-up period, has established LURIC as an important epidemiological resource from which multiple insights into CVD prevention and risk stratification have been derived.

The inclusion criteria for LURIC were as follows:

- Caucasian individuals of German ancestry residing in southwestern Germany;
- clinically stable at the time of enrollment;
- availability of a coronary angiogram; and
- provision of written informed consent.

All patient data have been **de-identified** prior to analysis; any patient records presented in this work (e.g., prompting examples) are synthetic and constructed solely for illustrative purposes.

Among the 3,316 patients included in the cohort, 2,600 (78.4%) were diagnosed with CVD, while follow-up data derived from previous studies (including CoroPredict [26] scores) are available for 2,112 (63.7%) of these CVD patients. Consequently, our analyses in Section 4 are conducted in two stages:

- the **full-cohort** of LURIC consisting of 3,316 patients; and
- the **sub-cohort** of LURIC consisting of 2,112 CVD patients with documented follow-up information.

#### Labels

Follow-up duration (in months) as well as the information about the death (binary) of a patient within the follow-up time is provided for all 3,316 patients in LURIC. One-year all-cause mortality (**1YM**) was therefore defined as “death occurring within 12 months of follow-up”, which we use as unified labels for all further analyses. Within the full-cohort, 139 (4.2%) patients died within 12 months of follow-up time from any cause, whereas in the CVD sub-cohort, 86 (4.1%) patients experienced mortality within the same period from any cause.

### 3.2 Selection of Biomarkers & Comorbidity Profiles

Our data preprocessing steps involved identifying and extracting five distinct sets of biomarkers and comorbidities as the underlying input for all regression and LLM analyses we conduct on LURIC.

#### 12 RiskyCAD Biomarkers (“Risk-12”)

RiskyCAD^2^ was an FP7 EU project (completed in 2017) which was developed to identify lipidomics and microRNA biomarkers associated with high-risk coronary atherosclerosis. These biomarkers, used alone or in conjunction with standard clinical risk models, aim to improve the detection of patients at elevated risk for myocardial infarction or cardiovascular mortality. All RiskyCAD biomarkers present in our dataset were identified and mapped to LURIC attributes, as summarized in Table 1.

**Table 1.** Collection of 12 RiskyCAD biomarkers stratified by 1-year mortality (1YM) over the full-cohort of LURIC [45]. Continuous variables are reported as *mean* ± *std*; categorical ones as *n* (%). Diabetes is mapped to *hba1c* and *dmwhoyn* in LURIC, while *supercrp, st2* and *fibrinog* in LURIC are summarized as *chronic inflammation* in the original RiskyCAD report.

|  |  | Mortality |  | P-Value |
| --- | --- | --- | --- | --- |
|  |  | 1YM=0 (*alive*) | 1YM=1 (*died*) |  |
| **Demographics** |  |  |  |  |
| *age* |  | 62.4 ± 10.6 | 68.6 ± 8.5 | <0.001 |
| *sex* | 1 | 2,206 (69.4%) | 104 (74.8%) | 0.209 |
|  | 2 | 971 (30.6%) | 35 (25.2%) |  |
| **Laboratory/Clinical** |  |  |  |  |
| *ldlch* |  | 116.8 ± 34.4 | 110.8 ± 32.9 | 0.038 |
| *hdlch* |  | 38.9 ± 10.8 | 34.6 ± 10.6 | <0.001 |
| *hba1c* |  | 6.3 ± 1.2 | 7.1 ± 1.7 | <0.001 |
| *cystatc* |  | 1.0 ± 0.4 | 1.4 ± 0.9 | <0.001 |
| *vitd25* |  | 17.6 ± 9.7 | 12.7 ± 7.4 | <0.001 |
| *pbnpl1* |  | 799.6 ± 1694.3 | 3306.1 ± 5466.1 | <0.001 |
| *tropt* |  | 1.3 ± 2.0 | 0.6 ± 1.1 | 0.017 |
| *Galectin3* |  | 15.5 ± 6.8 | 20.0 ± 9.2 | <0.001 |
| *supercrp* |  | 8.7 ± 17.7 | 17.1 ± 25.7 | <0.001 |
| *st2* |  | 21.6 ± 13.2 | 36.7 ± 43.3 | <0.001 |
| *fibrinog* |  | 393.7 ± 106.2 | 452.5 ± 111.9 | <0.001 |
| **Comorbidities** |  |  |  |  |
| *smoclass* | 0 | 1,148 (36.1%) | 46 (33.1%) | 0.152 |
|  | 1 | 1,396 (43.9%) | 72 (51.8%) |  |
|  | 2 | 633 (19.9%) | 21 (15.1%) |  |
| *dmwhoyn* | 0 | 2,280 (71.8%) | 61 (43.9%) | <0.001 |
|  | 1 | 897 (28.2%) | 78 (56.1%) |  |

#### 20 Core Biomarkers (“Core-20”)

A curated set of 20 routinely measured clinical biomarkers was compiled by medical experts for use as a baseline panel in this study. These biomarkers are listed in Table 2.

**Table 2.** Collection of 20 core biomarkers stratified by 1-year mortality (1YM) over the full-cohort of LURIC [45]. Continuous variables are reported as *mean* ± *std*; categorical ones as *n* (%). Creatinine is mapped to *crea* and *Crea*_*LCMSMS2* in LURIC, while total cholesterol *ech* extends *ldlch* and *hdlch*.

|  | Mortality |  | P-Value |
| --- | --- | --- | --- |
|  | 1YM=0 (*alive*) | 1YM=1 (*died*) |  |
| **Laboratory/Clinical** |  |  |  |
| *hb* | 13.9 ± 1.5 | 13.2 ± 1.7 | <0.001 |
| *leuco* | 7.1 ± 2.1 | 7.5 ± 2.5 | 0.059 |
| *avfgluc* | 112.7 ± 34.4 | 129.3 ± 51.1 | <0.001 |
| *hba1c* | 6.3 ± 1.2 | 7.1 ± 1.7 | <0.001 |
| *ech* | 192.9 ± 38.9 | 181.5 ± 40.9 | 0.002 |
| *ldlch* | 116.8 ± 34.4 | 110.8 ± 32.9 | 0.038 |
| *hdlch* | 38.9 ± 10.8 | 34.6 ± 10.6 | <0.001 |
| *etg* | 173.4 ± 118.8 | 164.9 ± 89.1 | 0.281 |
| *albumin* | 4.4 ± 0.6 | 4.2 ± 0.6 | <0.001 |
| *sgot* | 11.8 ± 7.7 | 12.6 ± 9.3 | 0.345 |
| *sgpt* | 16.7 ± 15.2 | 20.5 ± 60.1 | 0.455 |
| *ggt* | 26.6 ± 37.0 | 46.4 ± 100.4 | 0.022 |
| *crea* | 1.0 ± 0.3 | 1.2 ± 1.0 | 0.006 |
| *Crea_LCMSMS2* | 0.9 ± 0.5 | 1.1 ± 1.0 | 0.017 |
| *CKD_EPI_GFR* | 82.4 ± 19.7 | 67.7 ± 24.4 | <0.001 |
| *uricacid* | 5.1 ± 1.6 | 5.8 ± 2.7 | 0.004 |
| *potass* | 4.2 ± 0.3 | 4.1 ± 0.4 | 0.264 |
| *tsh* | 1.7 ± 4.2 | 1.5 ± 2.1 | 0.264 |
| *inrquick* | 1.1 ± 0.3 | 1.2 ± 0.4 | <0.001 |
| *aptt* | 37.5 ± 19.1 | 43.3 ± 25.4 | 0.009 |
| *supercrp* | 8.7 ± 17.7 | 17.1 ± 25.7 | <0.001 |
| *haptoglo* | 158.0 ± 74.9 | 179.6 ± 85.8 | 0.005 |

#### 21 Literature-derived Biomarkers (“Lit-20”)

An additional set of 21 clinically relevant biomarkers was identified through a targeted review of recent related literature on mortality analysis with LLMs [30, 38]. All biomarkers reported across these studies and available in our dataset were extracted for analysis. The complete list is presented in Table 3.

**Table 3.**
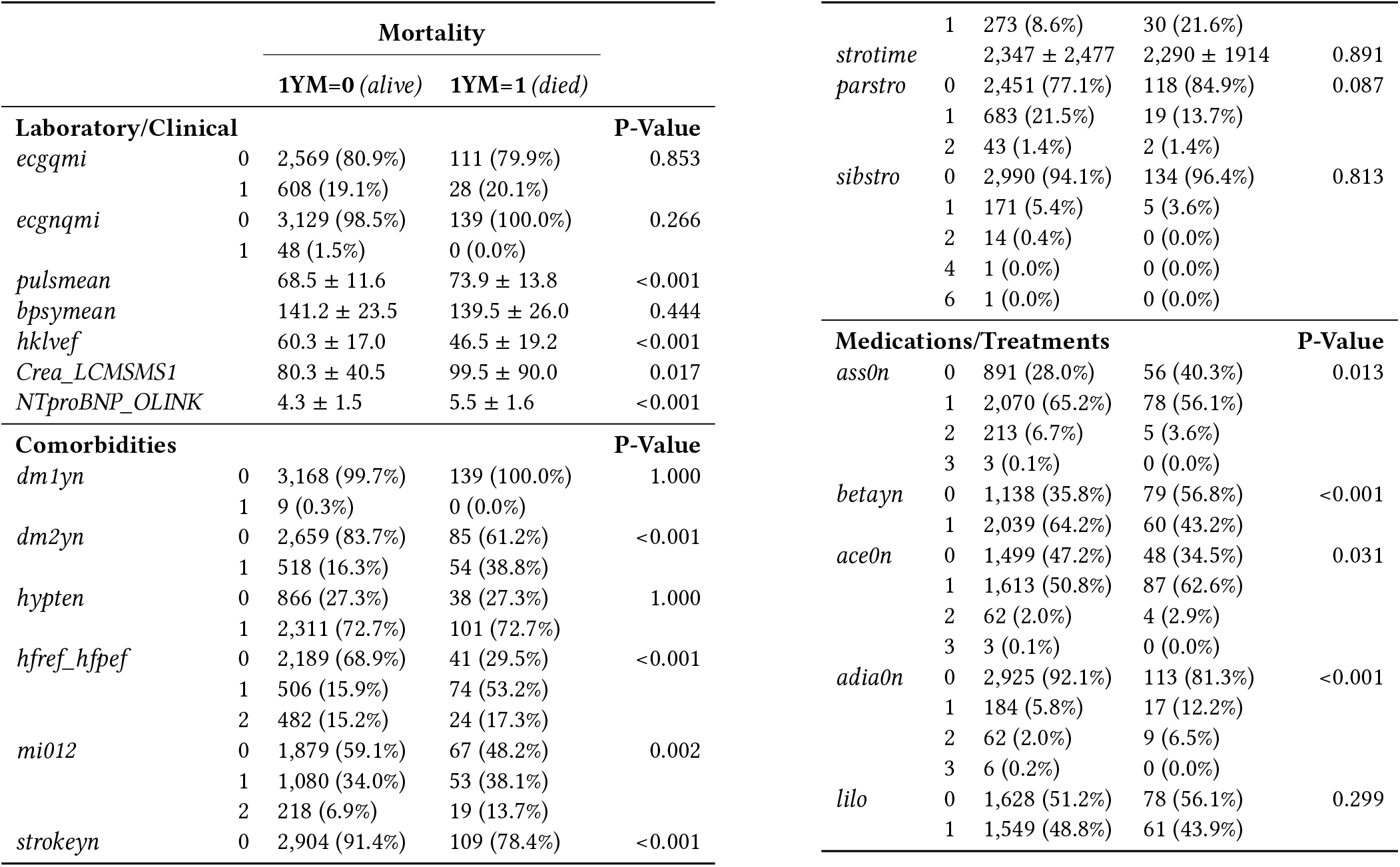
Collection of 21 literature-derived biomarkers stratified by 1-year mortality (1YM) over the full-cohort of LURIC [45]. Continuous variables are reported as *mean* ± *std*; categorical ones as *n* (%).

#### 64 Extended Biomarkers (“Ext-64”)

To broaden the scope of our analysis, we expanded the above biomarker sets through an extended literature review focused on predictors of all-cause mortality in patients with CVD [7, 26, 47]. This resulted in the 64 biomarkers documented in Table 4.

**Table 4.**
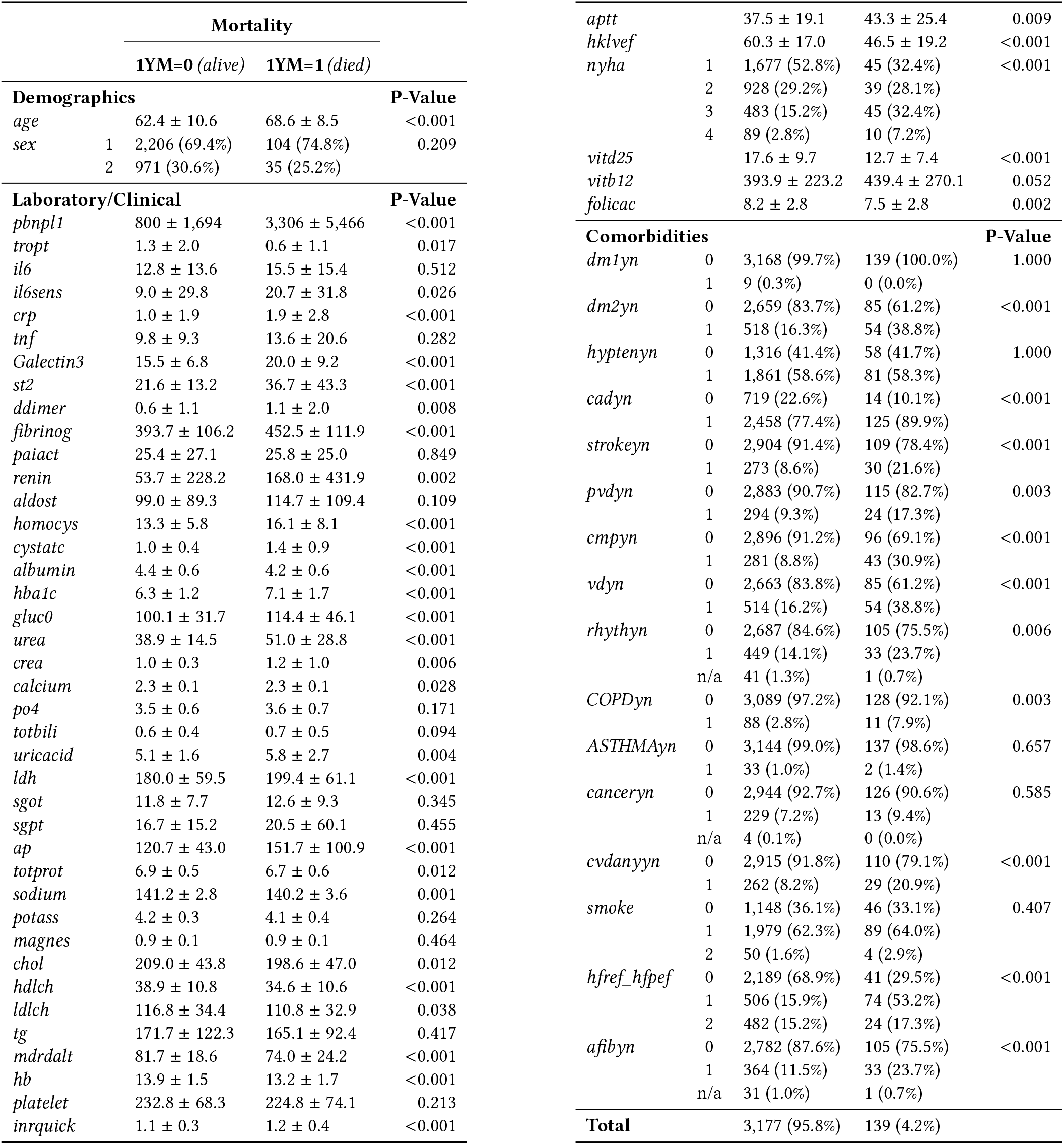
Extended collection of 64 biomarkers and comorbidities stratified by 1-year mortality (1YM) over the full-cohort of LURIC [45]. Continuous variables are reported as *mean* ± *std*; categorical ones as *n* (%).

#### 94 Comprehensive Biomarkers (“Comp-94”)

Finally, we created an aggregated biomarker set comprising 94 biomarkers by combining all of the four curated panels above. This comprehensive panel was used to evaluate the robustness of model performance across a broad biomarker spectrum.

A description of all biomarkers and comorbidities from the LURIC codebook is provided in Tables 7 to 10 in our Appendix. Attributes are depicted in the order in which the occur in the LURIC codebook. We remark that some of the markers from the literature map to multiple related attributes in LURIC (e.g., *crea, Crea_LCMSMS1* or *smoke, smoclass*). Other attributes in LURIC, like *supercrp, st2* and *fibrinogen*, have all been summarized under the term *chronic inflammation* in RiskyCAD. Moreover, patients’ general demographics (i.e., *age* and *sex*) have been included in all four sets for our experiments.

### 3.3 Regression: Boosting & Tabular Foundation Models

Gradient-boosted decision trees have remained the dominant methodology for modeling tabular data for more than two decades [18, 35]. However, with the expanding adoption of foundation models and ANNs for structured datasets, the need for standardized and reliable benchmarking frameworks has become increasingly critical. This gap has recently been addressed by the TabArena [12] initiative. Tabular Foundation Models (TFMs) are large, pretrained models specifically designed for tabular data. They aim to extend foundation-model capabilities—such as pretraining, transfer learning, and zero- or few-shot generalization—to structured datasets commonly used in many domains, including Finance, Healthcare, Marketing, and others. In this study, we focus on RealMLP^3^ [19] as a state-of-the-art representative of TFMs.

In addition, we evaluated the most widely used gradient-boosting algorithms, including LinearBoost^4^, CatBoost^5^ [35], and XGBoost^6^ [8], which continue to demonstrate strong performance in structured data applications. For additional context and baseline comparison, we also included Support Vector Machines^7^ (SVMs), a classic ML method extensively used in earlier predictive modeling research. A brief description of each algorithm is provided below:

- **LinearBoost** refers to boosting methods that use any type of base classifier—other than decision trees—as base learners. Each “weak” learner is a simple linear predictor, and boosting iteratively refines the ensemble by minimizing the loss function. This strategy preserves model linearity while improving predictive performance, making it particularly effective in high-dimensional and approximately linearly separable settings.
- **CatBoost** (short for “Categorical Boosting”) is a gradient boosting algorithm that natively handles categorical features, reducing or eliminating the need for one-hot encoding. It constructs an ensemble of decision trees trained sequentially, where each new tree corrects residual errors from previous iterations. CatBoost mitigates prediction shift and overfitting through ordered boosting and the use of symmetric tree structures.
- **XGBoost** (short for “eXtreme Gradient Boosting”) is a recent ML technique designed primarily for structured and tabular data, supporting both classification and regression tasks. Like CatBoost, it is also based on decision trees and uses gradient boosting over a series of trees. That is, it builds trees sequentially, where each new tree focuses on correcting the mistakes of the previous ones. XGBoost is efficient and consistently demonstrates strong performance in particular on tabular data.
- **RealMLP** denotes a real-valued Multi-Layer Perceptron (MLP), a neural network architecture designed to operate on continuous, real-valued input features. While structurally similar to standard MLPs, RealMLPs are emphasized in contexts where modeling continuous relationships is essential. They typically employ activation functions such as ReLU, *tanh*, or GELU to capture nonlinear interactions. Fully connected layers allow a dense information flow across the network. RealMLPs learn feature representations through backpropagation and are well suited for tasks such as regression, time-series modeling, tabular prediction, and general function approximation.
- **Support Vector Machines** (SVMs) denote a family of supervised learning algorithms used both for regression and classification. They aim to identify an optimal hyperplane that maximizes the margin between classes in the feature space. The approach relies on support vectors—the data points closest to the hyperplane. Nonlinear decision boundaries can be modeled through kernel functions (e.g., radial-basis functions and polynomial kernels). SVMs naturally support both regression and classification tasks by returning distances between the test vectors and the separating hyperplane.

#### Optimization

Each of the above regression techniques was optimized under a specific range of hyper-parameters using the Optuna^8^ framework with 200 trials per selection of cohort and biomarkers. The detailed ranges of hyperparameters considered for each method are depicted in Section B in our Appendix.

### 3.4 MedLLMs: Prompting Pipeline

Pretrained large language models (LLMs) are advanced neural network architectures—most commonly based on the Transformer framework—that learn statistical patterns of human language from massive text corpora. During pretraining, these models are exposed to trillions of tokens from diverse sources, enabling them to acquire general linguistic, semantic, and contextual representations without task-specific supervision. This foundational training equips LLMs with broad reasoning capabilities and the ability to generalize across domains.

LLMs are typically trained using self-supervised learning objectives, such as masked-token prediction or next-token prediction, which allow the model to infer structure and meaning from raw text. Once pretrained, the models can be adapted to downstream tasks—including classification, summarization, question answering, and clinical text interpretation—through finetuning or instruction prompting. This approach substantially reduces the need for large labeled datasets, making LLMs particularly valuable in fields where annotated data is limited or costly to obtain.

In biomedical and clinical applications, pretrained MedLLMs have demonstrated strong performance across tasks such as EHR analysis [25], clinical decision support [24], patient-triage automation [23], and extraction of structured information from unstructured notes [46]. Their ability to leverage broad linguistic knowledge while adapting to specialized medical contexts represents a significant advancement over traditional NLP approaches.

#### Zero-Shot Prompting

All biomarker sets (Section 3.2) were extracted from the tabular SPSS/SAV format of LURIC and then systematically converted into regularly structured textual prompts optimized for LLM comprehension. We added a fixed template header “*You are a medical expert analyzing patient data for mortality risk assessment. Based on the provided patient information, estimate the 1-year mortality risk of the patient as a percentage (0-100%). Consider all relevant clinical factors including age, sex, comorbidities, vital signs, laboratory values, and functional status*.”, which was then concatenated with the age and gender of the patient plus a list with the full names and units of the markers (e.g., “*ldlch*” is prompted as “*LDL cholesterol (EDTA) (mg/dL)*”) together with their respective values for the given patient (e.g., “*value of 118*”). We used “*value of*” in case of categorical or integer values and “*level of*” in case of continuous attributes with decimals (see Figure 2 for an example patient over the 12 RiskyCAD markers).

**Figure 2.**
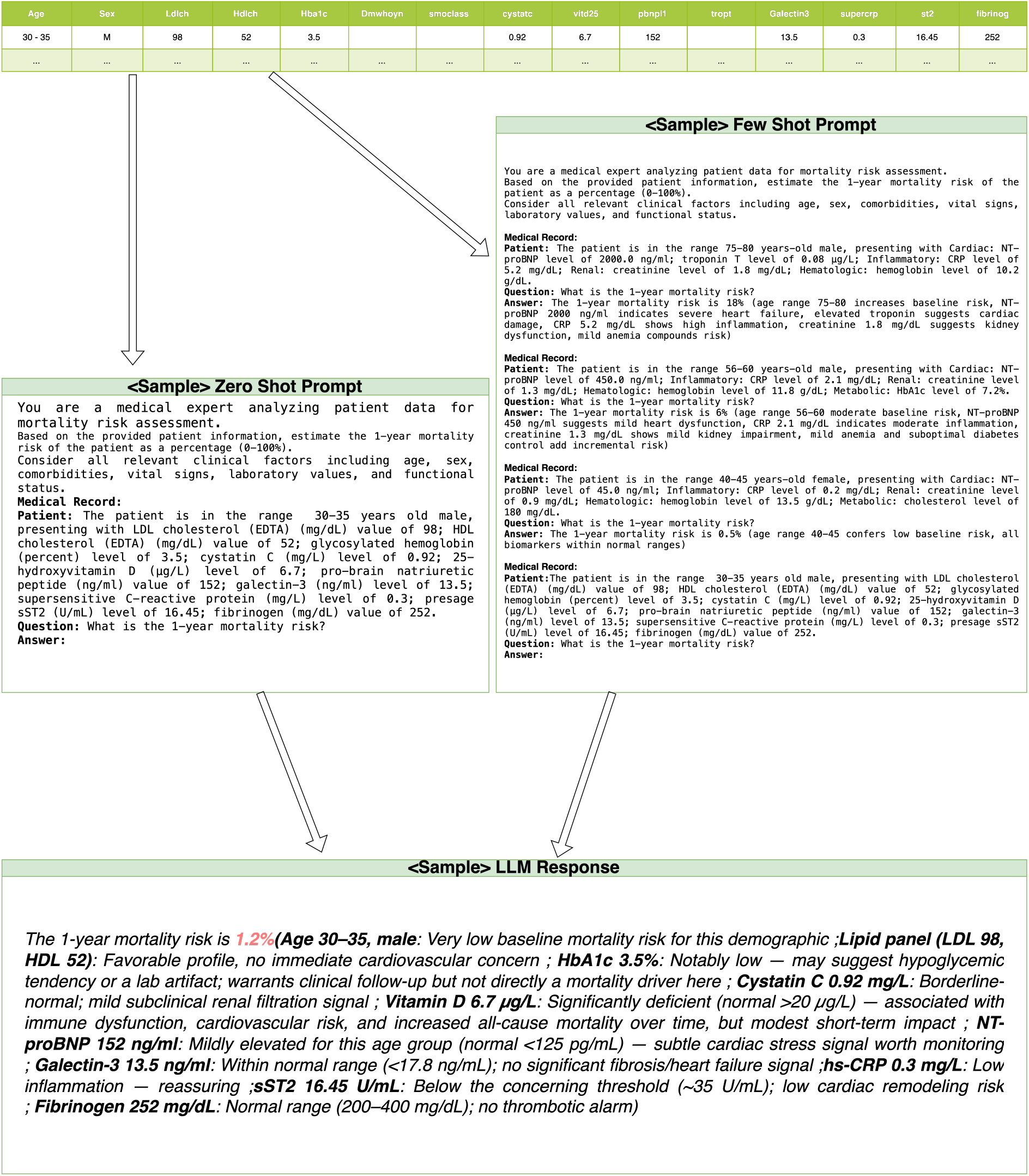
Prompting example (synthetic patient data) for RiskyCAD biomarkers (including *age* range, *sex*).

#### Few-Shot Prompting

We then extended our zero-shot prompting scheme to a few-shot scheme by prepending three synthethic records with given risk scores as shown in the illustrative example:

##### Patient

*The patient is in the range 75-80 years-old male, presenting with Cardiac: NT-proBNP level of 2000*.*0 ng/ml; troponin T level of 0*.*08 µg/L; Inflammatory: CRP level of 5*.*2 mg/dL; Renal: creatinine level of 1*.*8 mg/dL; Hematologic: hemoglobin level of 10*.*2 g/dL*.

##### Question

*What is the 1-year mortality risk?*

##### Answer

*The 1-year mortality risk is 18% (age range 75-80 increases baseline risk, NT-proBNP 2000 ng/ml indicates severe heart failure, elevated troponin suggests cardiac damage, CRP 5*.*2 mg/dL shows high inflammation, creatinine 1*.*8 mg/dL suggests kidney dysfunction, mild anemia compounds risk)*

##### Patient

*The patient is in the range 56-60 years-old male, presenting with Cardiac: NT-proBNP level of 450*.*0 ng/ml; Inflammatory: CRP level of 2*.*1 mg/dL; Renal: creatinine level of 1*.*3 mg/dL; Hematologic: hemoglobin level of 11*.*8 g/dL; Metabolic: HbA1c level of 7*.*2%*.

##### Question

*What is the 1-year mortality risk?*

##### Answer

*The 1-year mortality risk is 6% (age range 56-60 moderate baseline risk, NT-proBNP 450 ng/ml suggests mild heart dysfunction, CRP 2*.*1 mg/dL indicates moderate inflammation, creatinine 1*.*3 mg/dL shows mild kidney impairment, mild anemia and suboptimal diabetes control add incremental risk)*

##### Patient

*The patient is in the range 40-45 years-old female, presenting with Cardiac: NT-proBNP level of 45*.*0 ng/ml; Inflammatory: CRP level of 0*.*2 mg/dL; Renal: creatinine level of 0*.*9 mg/dL; Hematologic: hemoglobin level of 13*.*5 g/dL; Metabolic: cholesterol level of 180 mg/dL*. ***Question:*** *What is the 1-year mortality risk?*

##### Answer

*The 1-year mortality risk is 0*.*5% (age range 40-45 confers low baseline risk, all biomarkers within normal ranges)*

The translated patient records were then sent to the LLMs for the generation of the desired answers. Both the structure and the content of our few-shot template triggered all LLMs we investigated in our experiments (Section 3.5) to generate numeric risk scores within the first lines of their answers in a remarkably reliable manner. We thus used a regular expression to extract the generated mortality risk scores from the outputs (also shown in Figure 2).The prompting scheme illustrated in Figure 2 uses synthetic patient data for demonstration purposes only.

### 3.5 Choice of Models

For both prompting schemes, we investigated a wide range of openly available MedLLMs from the HuggingFace Medical LLM leaderboard, including MedGemma-27B^9^, Med42-70B^10^, MMed70B^11^, MedLlama-8B^12^, meditron-70b^13^ and OpenBioLLM-70B^14^, and we also included Llama-3.1-70B^15^ (and its variants) as baseline. The `AutoModelForCausalLM` class of the Transformers Python API was used for prompting MedGemma and the Llama-based models. To provide a comprehensive comparison, we also incorporated three state-of-the-art commercial models, namely Claude Sonnet 4.5, ChatGPT 5.2 and Gemini 3, via their proprietary Python APIs.

### 3.6 MedLLMs: Supervised Finetuning

In addition to prompting, we also implemented a supervised finetuning workflow that further trains the above MedLLMs directly on our zero-shot prompts. This approach allows the model to learn clinically relevant patterns while preserving its ability to perform natural-language inference. Given the substantial computational requirements, finetuning was limited to the 4B parameter version of MedGemma and the 8B variants of the Llama-based models, respectively. We use the `AutoModelForSequenceClassification` API of Transformers for classification finetuning by training a LoRa [20] adapter on top of both MedGemma and the Llama-based models, and by using the 1YM labels (Section 3.1) as targets.

## 4 Experiments

To rigorously evaluate the proposed models and assess their effectiveness in predicting 1-year all-cause mortality in the two cohorts of LURIC described in Section 3.1, we conducted a comprehensive series of experiments combining classic Machine Learning (ML) algorithms, Tabular Foundation Models (TFMs), and Large Language Models (LLMs). All experiments were conducted on a dual **H100 NVLink** platform (running CUDA Version 13.0) with the following configuration.

- **CPU cores:** 48 (4 × Intel(R) Xeon(R) 6505P)
- **Main memory:** 256 GB DDR2
- **Interface:** PCIe 5.0
- **CUDA cores:** 33,792 (2 × Nvidia H100)
- **GPU memory:** 192 GB HBM3
- **Tensor cores:** 1,056 (4^*th*^-generation architecture)
- **Interconnect:** 18 × (4^*th*^-generation NVLink)
- **Maximum FP8 performance:** 3,958 TFLOPS

All models were evaluated under standardized experimental conditions using harmonized input features derived from the clinical biomarkers and comorbidity profiles described in Section 3.2. We assessed the various LLMs’ performances under zero- and few-shot prompting settings, followed by supervised finetuning of the openly available LLMs. The following subsections detail the experimental configurations, training procedures, and evaluation criteria used throughout this study.

### 4.1 Experimental Configurations of Machine Learning & Tabular Foundation Models

To address the substantial class imbalance in the dataset, all ML models (SVM, LinearBoost, CatBoost & XGBoost)—including the Tabular Foundation Model (RealMLP)—were evaluated using 10-fold cross-validations under each of the 200 Optuna trials per regressor, cohort, and biomarker selection. Overall, this resulted in several hundreds of thousands of individual runs just for the regression part of our experiments, taking about one week on the above server configuration to finish. This strategy however provides a robust and reliable estimate of model performance by reducing sensitivity to sampling variability and hyperparameters.

### 4.2 Experimental Configurations of LLM Prompting

In contrast, the prompting-based LLM experiments were not cross-validated as they did not involve any training/testing steps. Rather, we performed direct in-context learning demonstrations with three examples (“3-shots”) for each run. This decision was made to prevent potential information leakage across folds, avoid unintended retention of prior context by the LLM, and mitigate constraints related to model memory and context window limitations. We deployed a distributed generation pipeline by using the latest vLLM 0.18.4^16^ API which provides and efficient wrapper for the Transformer API from HuggingFace. These prompting experiments took about one day to finish on our GPU server configuration.

### 4.3 Experimental Configurations of LLM Classification Finetuning

Given the substantial computational requirements, finetuning experiments were limited to 5-fold cross-validations, and we also reduced our tests to the 4B parameter version of MedGemma and the 8B variants of the Llama-based models, respectively. DeepSpeed 0.18.4^17^ was deployed on top of our Transformers implementation for accelerated distributed training by using a batch size of 16, 8 training epochs, and a learning rate of 1e-4 with Cosine scheduling. The finetuning of all runs took about two days on our GPU server to finish.

### 4.4 Evaluation Criteria

Accuracy alone is an insufficient indicator of model performance in mortality prediction tasks, largely due to the inherent class imbalance in these datasets. Prior studies have demonstrated that certain large language models (LLMs) may exhibit deceptively high accuracy by correctly identifying the majority of survivors (true negatives) while failing to recognize a substantial proportion of non-survivors (false negatives) [38]. Given this imbalance, the *Area Under the Receiver Operating Characteristic* (AUROC) curve serves as a more appropriate metric for assessing a model’s discriminative ability, which is also consistent with previous evaluations [6, 41, 42].

In clinical settings, however, strong discrimination is not enough; reliable probability estimates are also required. Therefore, we additionally assess model calibration using the *Expected Calibration Error* (ECE) [14, 34]. Across evaluations, LLM-based models tended to underestimate mortality risk, underscoring the need for post-hoc calibration techniques to adjust predicted probabilities and enhance their clinical trustworthiness.

Model performance was thus evaluated using ROC-based rankings and ECE-based calibration curves. The area under the ROC curve (AUROC) quantified the discriminative ability for 1-year mortality prediction. Calibration was assessed by comparing predicted probabilities to observed event rates across probability bins. To address systematic miscalibration, we applied Platt scaling—a post-hoc method that fits a logistic regression on top of the initial model predictions in order to map these to calibrated probabilities.

### 4.5 Detailed AUROC Results

Tables 5 and 6 depict our detailed AUROC results for the two cohorts of LURIC (Section 3.1) in combination with our five biomarker sets (Section 3.2). Selected ROC plots are depicted in Figure 3 for the sub-cohort of LURIC. We find that commercially available LLMs, when combined with appropriate prompting strategies, achieve strong discriminative performance, with AUROC values of up to 0.820 on the full-cohort of LURIC and up to 0.849 on the sub-cohort (Gemini-3-Flash), both under the Ext-64 marker set. These results are comparable to the best regression model (CatBoost) in our setting, which also achieves an AUROC of 0.849 on the same sub-cohort and marker set. Smaller models (8B), on the other hand, can be finetuned to meet or even exceed their larger (70B) counterparts, as it is, e.g., the case for Meditron3-8B with an excellent AUC value of 0.826 on the full-cohort of LURIC and the Ext-64 marker set. We remark that the provided CoroPredict [26] scores over the sub-cohort yield an AUROC of 0.841, while SMART [39] and SCORE2 [37] yield 0.729 and 0.668, respectively (these are however available only for different cohorts of LURIC).

**Table 5.** Detailed AUROC results for the full-cohort of LURIC.

| Method/<br>Model | Biomarker Collection |  |  |  |  |
| --- | --- | --- | --- | --- | --- |
|  | Risk-12 | Base-20 | Lit-21 | Ext-64 | Comp-94 |
| REGRESSION: BOOSTING & TABULAR FOUNDATION MODELS |  |  |  |  |  |
| SVM | 0.817 (± 0.059) | 0.722 (± 0.047) | 0.765 (± 0.044) | 0.819 (± 0.062) | 0.823 (± 0.058) |
| LinearBoost | 0.826 (± 0.051) | 0.776 (± 0.050) | 0.795 (± 0.057) | 0.826 (± 0.042) | 0.826 (± 0.052) |
| CatBoost | 0.829 (± 0.052) | **0.792** (± 0.049) | **0.816** (± 0.047) | **0.839** (± 0.051) | **0.841** (± 0.053) |
| XGBoost | 0.829 (± 0.052) | 0.780 (± 0.057) | 0.810 (± 0.044) | 0.825 (± 0.055) | 0.828 (± 0.050) |
| RealMLP | **0.830** (± 0.017) | 0.777 (± 0.024) | 0.813 (± 0.013) | 0.836 (± 0.023) | 0.839 (± 0.021) |
| FEW-SHOT PROMPTING (3-SHOT PROMPTS) |  |  |  |  |  |
| MedGemma-27B | 0.762 | 0.698 | 0.698 | 0.762 | 0.749 |
| Med42-70B | **0.796** | **0.727** | 0.649 | **0.815** | **0.786** |
| MMed-70B | 0.770 | 0.609 | 0.624 | 0.806 | 0.714 |
| JSL-MedLlama-8B | 0.726 | 0.652 | 0.715 | 0.785 | 0.754 |
| meditron-70b | 0.728 | 0.632 | 0.687 | 0.779 | 0.716 |
| OpenBioLLM-70B | 0.731 | 0.698 | **0.722** | 0.777 | 0.743 |
| Llama-3-8B | 0.722 | 0.653 | 0.682 | 0.768 | 0.543 |
| Llama-3-70B | 0.688 | 0.630 | 0.678 | 0.715 | 0.679 |
| Llama-3-8B-Inst | 0.736 | 0.666 | **0.715** | 0.789 | **0.776** |
| Llama-3-70B-Inst | **0.778** | **0.727** | 0.713 | **0.798** | 0.759 |
| Llama-3.1-8B | 0.716 | 0.579 | 0.675 | 0.690 | 0.649 |
| Llama-3.1-70B | 0.741 | 0.641 | **0.707** | 0.765 | 0.723 |
| Llama-3.1-8B-Inst | 0.727 | 0.681 | 0.676 | **0.812** | **0.771** |
| Llama-3.1-70B-Inst | **0.766** | **0.700** | 0.684 | 0.788 | 0.758 |
| Gemini-3-Flash | **0.804** | 0.766 | 0.744 | **0.820** | **0.812** |
| Claude-Sonnet-4.5 | 0.795 | 0.767 | **0.749** | 0.815 | 0.810 |
| ChatGPT-5.2 | 0.710 | **0.772** | 0.672 | 0.808 | 0.800 |
| CLASSIFICATION FINETUNING (0-SHOT PROMPTS) |  |  |  |  |  |
| MedGemma-4B | 0.740 (± 0.020) | 0.654 (± 0.059) | 0.772 (± 0.027) | 0.814 (± 0.021) | 0.750 (± 0.056) |
| Med42-8B | 0.783 (± 0.040) | **0.697** (± 0.050) | 0.718 (± 0.034) | 0.816 (± 0.031) | 0.743 (± 0.052) |
| MMed-8B | 0.780 (± 0.040) | 0.668 (± 0.031) | 0.771 (± 0.048) | 0.815 (± 0.018) | 0.805 (± 0.050) |
| JSL-MedLlama-8B | 0.782 (± 0.041) | 0.682 (± 0.077) | 0.766 (± 0.022) | 0.808 (± 0.033) | 0.726 (± 0.080) |
| Meditron3-8B | **0.813** (± 0.045) | 0.684 (± 0.087) | **0.784** (± 0.019) | **0.826** (± 0.025) | **0.809** (± 0.028) |
| OpenBioLLM-8B | 0.784 (± 0.052) | 0.686 (± 0.028) | 0.749 (± 0.041) | 0.810 (± 0.027) | 0.773 (± 0.035) |
| Llama-3.1-8B | 0.778 (± 0.048) | 0.658 (± 0.063) | 0.706 (± 0.071) | 0.824 (± 0.026) | 0.753 (± 0.059) |

**Table 6.**
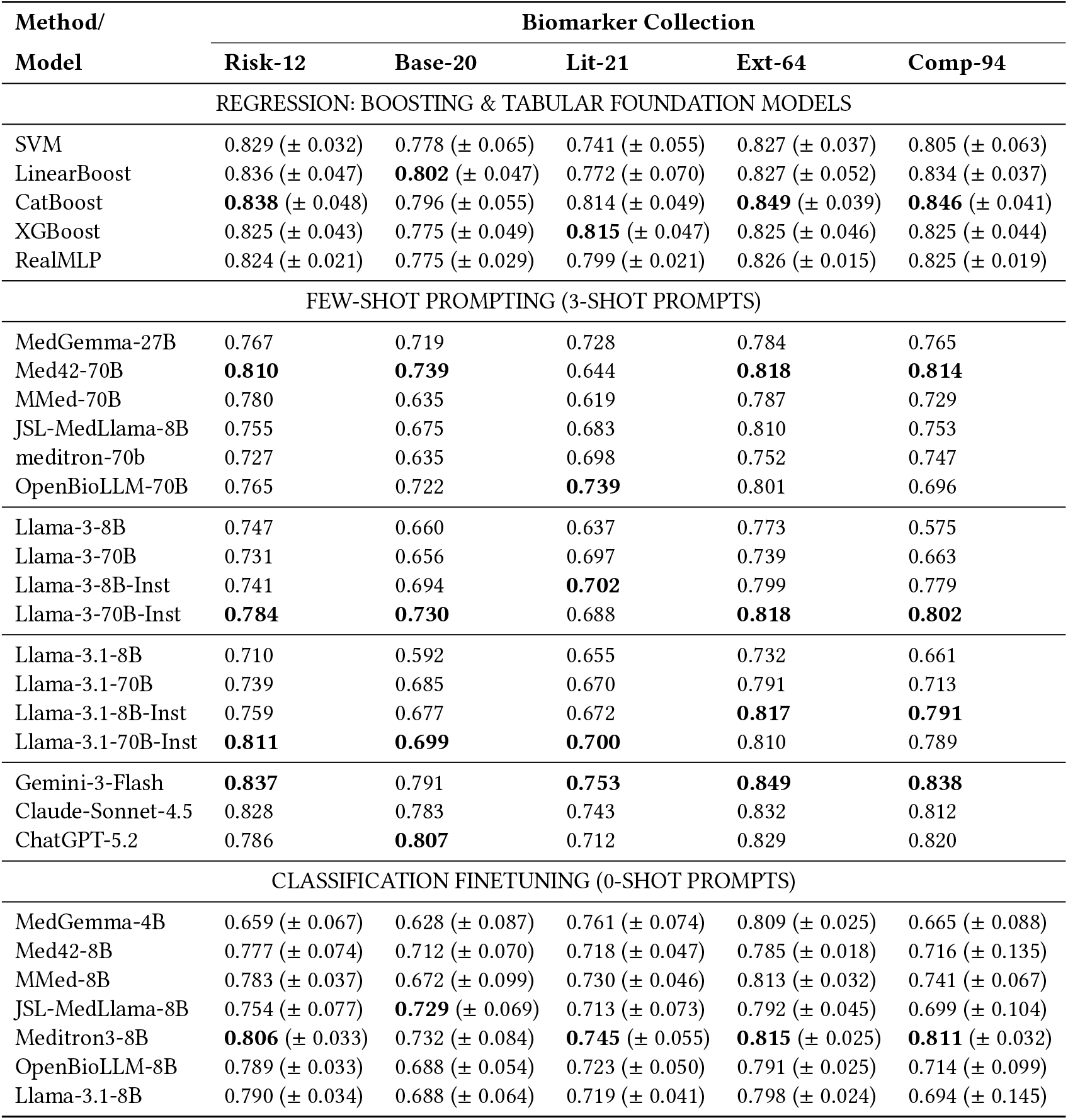
Detailed AUROC results for the sub-cohort of LURIC.

**Figure 3.**
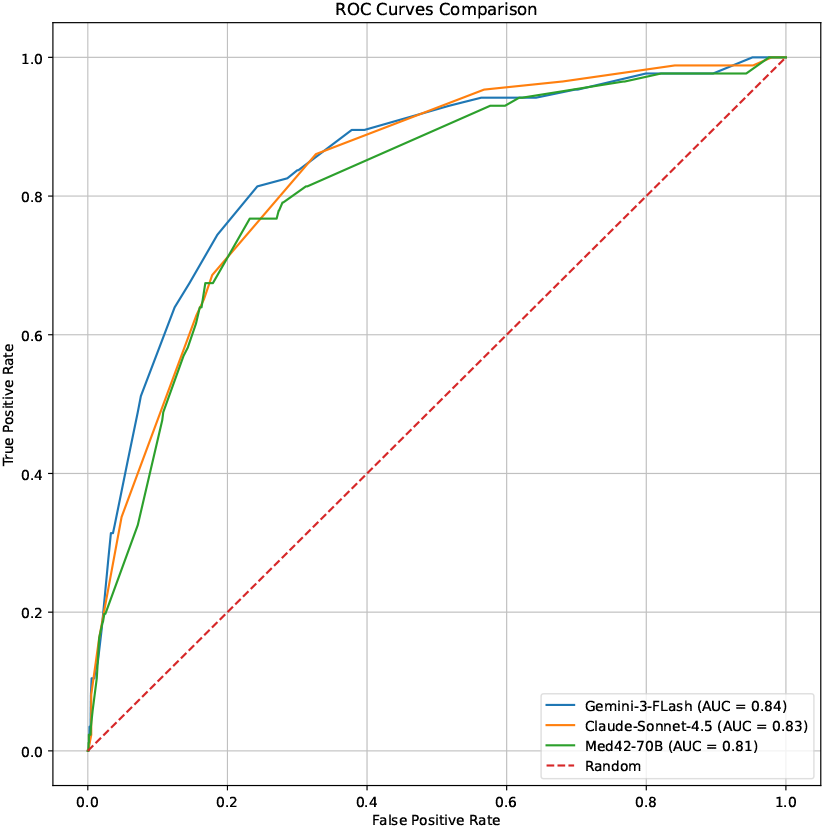
Selected ROC curves over the sub-cohort of LURIC and the Ext-64 markers.

### 4.6 Model Stratification & Calibration

We further observe that LLMs systematically under-predict mortality risk when prompted to generate risk scores. This miscalibration can be substantially mitigated through post-hoc calibration techniques, such as Platt scaling [34]. As illustrated for Med42-70B in Figure 4, calibration reduces the *Expected Calibration Error* (ECE) metric by up to 90%. Among the evaluated models, commercial models such as Gemini-3-Flash demonstrated the lowest ECE, indicating a comparatively better inherent calibration.

**Figure 4.**
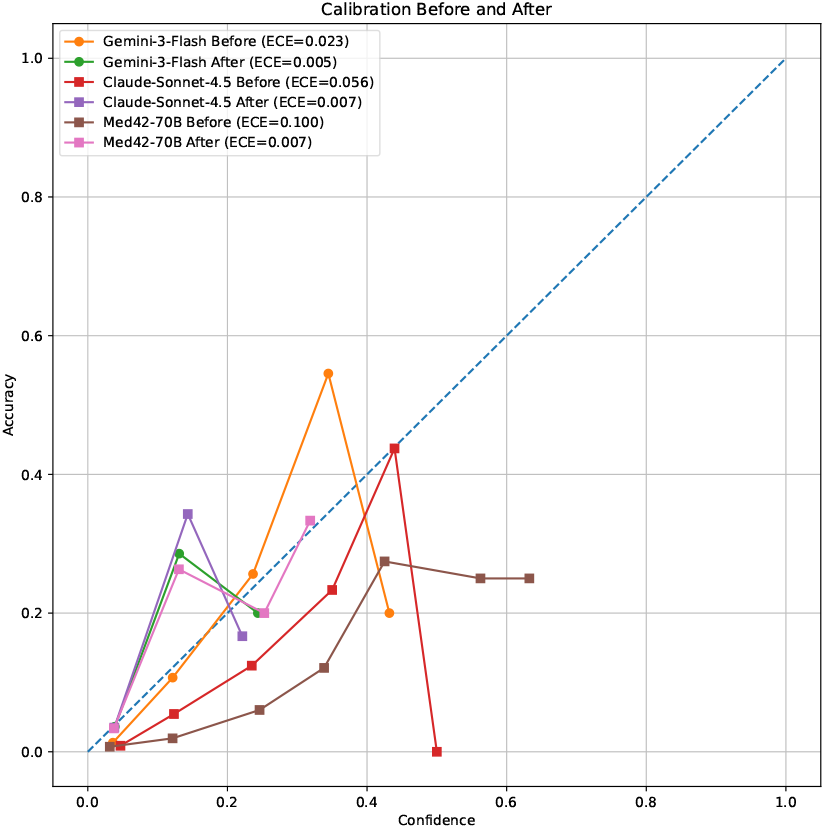
Selected calibration plots over the sub-cohort of LURIC and the Ext-64 markers.

## 5 Conclusions & Outlook

Our study demonstrates the strong potential of MedLLMs for predicting 1-year all-cause mortality in CVD patients under a wide range of routinely collected biomarkers. Our key findings include that (a) large, pretrained MedLLMs (70B) achieve up to 82% AUROC with optimized few-shot prompting, thus making them competitive to the best regression-based(CatBoost) and state-of-the-art techniques in the medical literature such as CoroPredict (both ranging at 84% AUROC under the sub-cohort of LURIC); (b) smaller MedLLMs (8B) can be finetuned to match or even surpass both their larger counterparts as well as commercial models such as ChatGPT-5.2 and Claude-Sonnet-4.5 (exceeding 82% AUROC); (c) further model-calibration and -stratification analyses reveal a systematic mortality over-prediction (ECE: 0.05–0.10) of MedLLMs, while Platt scaling effectively reduces miscalibrations by 60–90%. Our study reveals large gains of recent LLMs for CVD and mortality predictions compared to earlier results [1, 5, 30], where LLMs still showed a significant gap to leading regression techniques, ANNs, and the CoroPredict baseline. A particularly remarkable result however is the strong performance of Gemini-3-Flash, which is on par with our best regression technique (CatBoost) and the CoroPredict baseline just by few-shot prompting—without any domain-specific adaptations or finetuning, which also demonstrates a significant gain over prior prompting results [15, 49].

Overall, our findings demonstrate that MedLLMs, when properly prompted and finetuned using routinely available and cost-effective biomarkers, can function as highly accurate and interpretable tools for clinical risk prediction. As such, they may be used either directly on top of tabular data (such as EHRs) or also in combination with further textual notes (e.g., clinical notes or discharge records). As future work, we plan to further explore finetuning techniques based on Reinforcement Learning from Human Feedback (RLHF), as well as to further optimize our biomarker selection under a budget-constrained optimization framework.

## Data Availability

This study made use of the LURIC (LUDWIGSHAFEN RISK AND CARDIOVASCULAR HEALTH STUDY) dataset, which can be accessed upon reasonable request at https://www.luric.online/luric2.html

## Appendix A Biomarker & Comorbidity Statistics

**Table 7.** Risk-12 Markers.

| Marker | Description (Unit) |
| --- | --- |
| **Demographics** |  |
| age | Patient’s age (years) |
| sex | 1=male, 2=female |
| **Laboratory/Clinical** |  |
| ldlch | LDL cholesterol (EDTA) (mg/dL) |
| hdlch | HDL cholesterol (EDTA) (mg/dL) |
| hba1c | Glycosylated hemoglobin (%) |
| cystatc | Cystatin C (mg/L) |
| vitd25 | 25-hydroxy vitamin D (µg/L) |
| pbnpl1 | Pro-brain natriuretic peptide (ng/mL) |
| tropt | Troponin T (µg/L) |
| Galectin3 | Galectin-3 (ng/mL) |
| supercrp | Supersensitive C-reactive protein (mg/L) |
| st2 | Presage sST2 (U/mL) |
| fibrinog | Fibrinogen (mg/dL) |
| **Comorbidities** |  |
| smoclass | Smoking class (0=no, 1=quit>30days, 2=active) |
| dmwhoyn | Diabetes status (WHO criteria) |

**Table 8.** Core-20 Markers.

| Marker | Description (Unit) |
| --- | --- |
| **Laboratory/Clinical** |  |
| hb | Hemoglobin (g/dL) |
| leuco | Leucocytes (/nL) |
| avfgluc | Mean fasting glucose (serum/plasma) (mg/dL) |
| hba1c | Glycosylated hemoglobin (%) |
| ech | Total cholesterol (EDTA) (mg/dL) |
| ldlch | LDL cholesterol (EDTA) (mg/dL) |
| hdlch | HDL cholesterol (EDTA) (mg/dL) |
| etg | Triglycerides (EDTA) (mg/dL) |
| albumin | Albumin (g/dL) |
| sgot | SGOT (U/L) |
| sgpt | SGPT (U/L) |
| ggt | Gamma GT (U/L) |
| crea | Creatinine (mg/dL, Jaffe) |
| Crea_LCMSMS2 | Creatinine (mg/dL) |
| CKD_EPI_GFR | Chronic kidney disease epidemiology coll. |
| uricacid | Uric acid (mg/dL) |
| potass | Potassium (mmol/L) |
| tsh | TSH (mU/L) |
| inrquick | INR Quick (%) |
| aptt | aPTT (sec) |
| supercrp | Supersensitive C-reactive protein (mg/L) |
| haptoglo | Haptoglobin polymorphism |

**Table 9.** Base-21 Markers.

| Marker | Description (Unit) |
| --- | --- |
| **Laboratory/Clinical** |  |
| ecgqmi | ST elevation (Q wave) MI (cath. report) |
| ecgnqmi | Non-ST elevation (non Q) MI (cath. report) |
| pulsmean | Mean heart rate (beats per minute) |
| bpsymean | Mean systolic blood pressure (mmHg) |
| hklvef | Angiographic ejection fraction (cath. report) |
| Crea_LCMSMS1 | Creatinine (µmol/L) |
| NTproBNP_OLINK | N-terminal prohormone (NT-proBNP) |
| **Comorbidities** |  |
| dm1yn | Type I diabetes mellitus |
| dm2yn | Type II diabetes mellitus |
| hypten | Hypertension |
| hfref_hfpef | Type of heart failure |
| mi012 | Myocardial infarction |
| strokeyn | Stroke/PRIND/TIA |
| strotime | Time since first stroke (days) |
| parstro | Number of parents having stroke (0-2) |
| sibstro | Number of siblings having stroke (0-N) |
| **Medications/Treatments** |  |
| ass0n | Aspirin/other antiplatelet |
| betayn | Beta blocker |
| ace0n | ACE-inhibitor (0-N) |
| adia0n | Oral antidiabetic agent (0-N) |
| lilo | Lipid lowering therapy |

**Table 10.** Ext-64 Markers.

| Marker | Description (Unit) |  |  |
| --- | --- | --- | --- |
| **Demographics** |  |  |  |
| *age* | Patient's age (years) | *potass* | Potassium (mmol/L) |
| *sex* | 1=male, 2=female | *magnes* | Magnesium (mmol/L) |
| **Laboratory/Clinical** |  | *chol* | Cholesterol (serum) (mg/dL) |
| *pbnpl1* | Pro-brain natriuretic peptide (ng/mL) | *ldlch* | LDL-cholesterol (serum) (mg/dL) |
| *tropt* | Troponin T (μg/LF) | *hdlch* | HDL-cholesterol (serum) (mg/dL) |
| *il6* | Interleukin 6 (ng/L) | *tg* | Triglycerides (serum) (mg/dL) |
| *il6sens* | IL-6 sensitive assay (ng/L) | *mdrdalt* | eGFR MDRD Crea (Jaffe) |
| *crp* | C-reactive protein (mg/dL) | *hb* | Hemoglobin (g/dL) |
| *tnf* | Tumor necrosis factor (TNF) a (ng/L) | *platelet* | Platelets (/nL) |
| *Galectin3* | Galectin-3 (ng/mL) | *inrquick* | INR Quick (%) |
| *st2* | Presage sST2 (U/mL) | *aptt* | aPTT (sec) |
| *ddimer* | D-dimer (mg/L) | *hklvef* | Angiographic ejection fraction (cath. report) |
| *fibrinog* | Fibrinogen (mg/dL) | *nyha* | NYHA heart failure classification (1-4) |
| *paiact* | PAI-1 activity (U/mL) | *vitb12* | Vitamin B12 (ng/L) |
| *renin* | Renin (U/L) | *vitd25* | 25-hydroxy vitamin D (μg/L) |
| *aldost* | Aldosterone (ng/L) | *folicac* | Folic acid (μg/L) |
| *homocys* | Homocysteine (μmol/L) | **Comorbidities** |  |
| *cystatc* | Cystatin C (mg/L) | *dm1yn* | Type I diabetes mellitus |
| *albumin* | Albumin (g/dL) | *dm2yn* | Type II diabetes mellitus |
| *hba1c* | Glycosylated hemoglobin (%) | *hyptenyn* | History of arterial hypertension |
| *gluc0* | Fasting glucose (whole blood) (mg/dL) | *cadyn* | CAD (>10% (=20% or more)) or clinical |
| *urea* | Urea (mg/dL) | *strokeyn* | Stroke/PRIND/TIA |
| *crea* | Creatinine (mg/dL, Jaffe) | *pvdyn* | Peripheral vascular disease |
| *calcium* | Calcium (mmol/L) | *cmpyn* | Cardiomyopathy |
| *po4* | Anorganic phosphate (mg/dL) | *vdyn* | Vascular disease |
| *totbili* | Total bilirubin (mg/dL) | *rhythyn* | Arrhythmia |
| *uricacid* | Uricacid (mg/dL) | *COPDyn* | Chronic obstructive pulmonary disease |
| *ldh* | LDH (U/L) | *ASTHMAyn* | Asthma |
| *sgot* | SGOT (U/L) | *canceryn* | Active cancer (any type) |
| *sgpt* | SGPT (U/L) | *cvdanyyn* | Cerebrovascular (incl. atherosclerotic) disease |
| *ap* | Alkaline phosphatase (U/L) | *smoke* | Smoker (0=never, 1=cigarette, 2=pipe/cigar) |
| *totprot* | Total protein (g/dL) | *hfref_hfpef* | Type of heart failure |
| *sodium* | Sodium (mmol/L) | *afibyn* | Atrial fibrillation |

## B Hyperparameters Considered for Optimization

**Listing 1.**
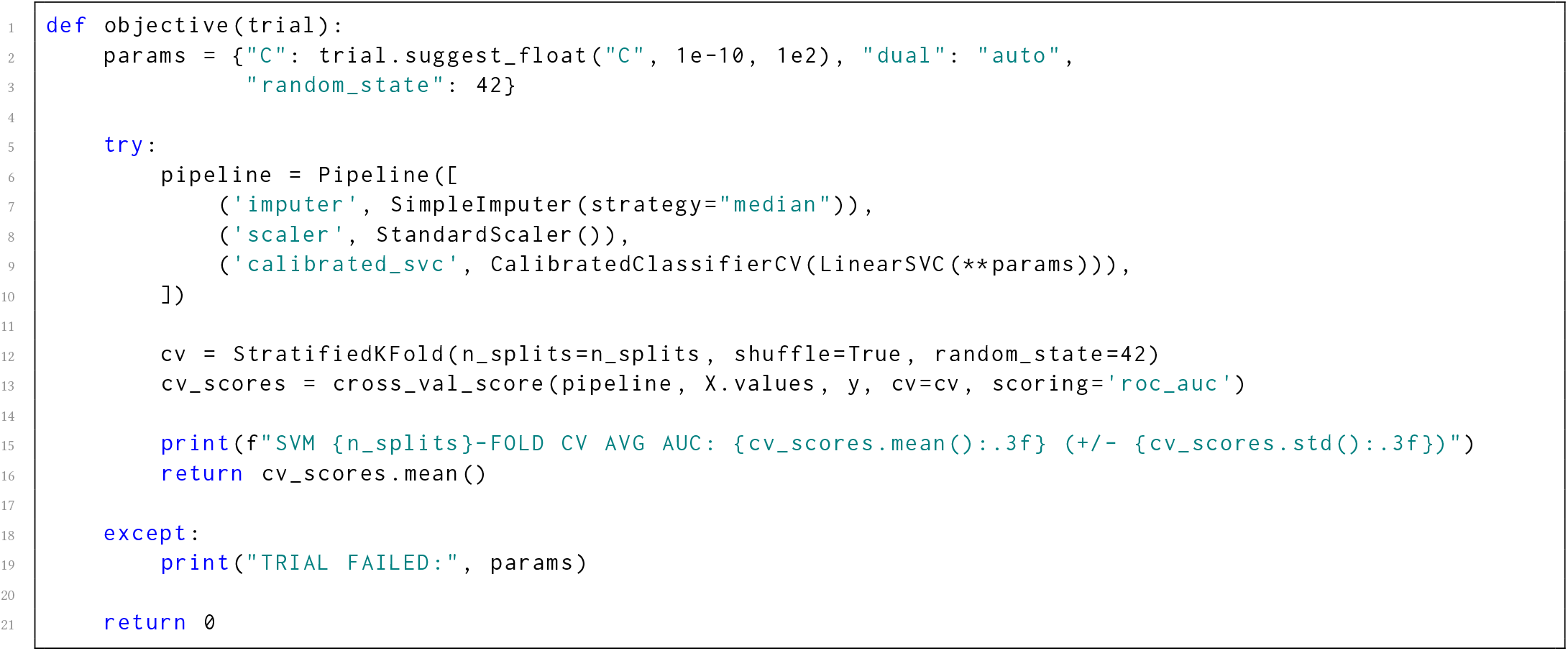
Optuna Hyperparameters for SVM/SVC.

**Listing 2.**
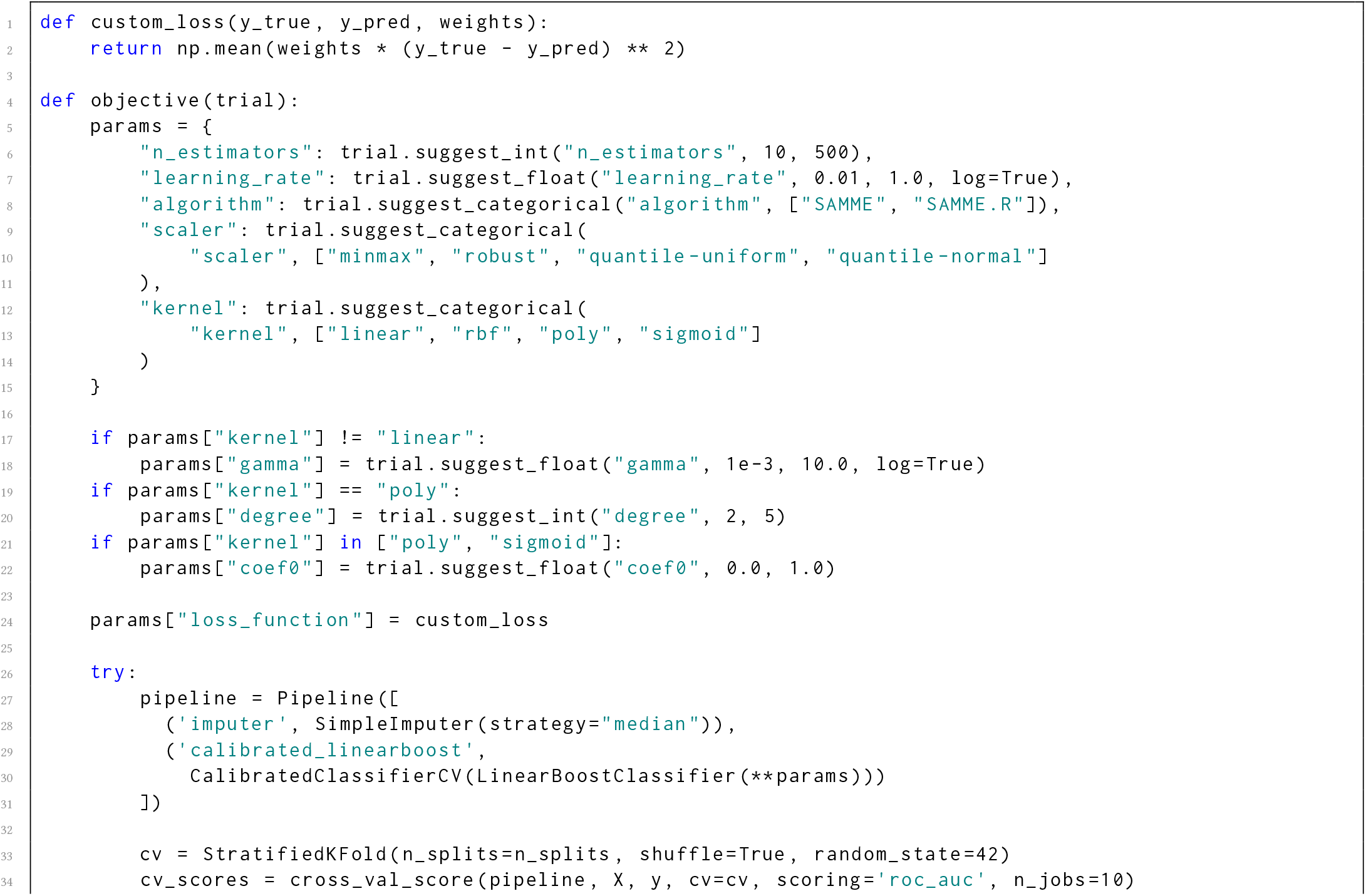
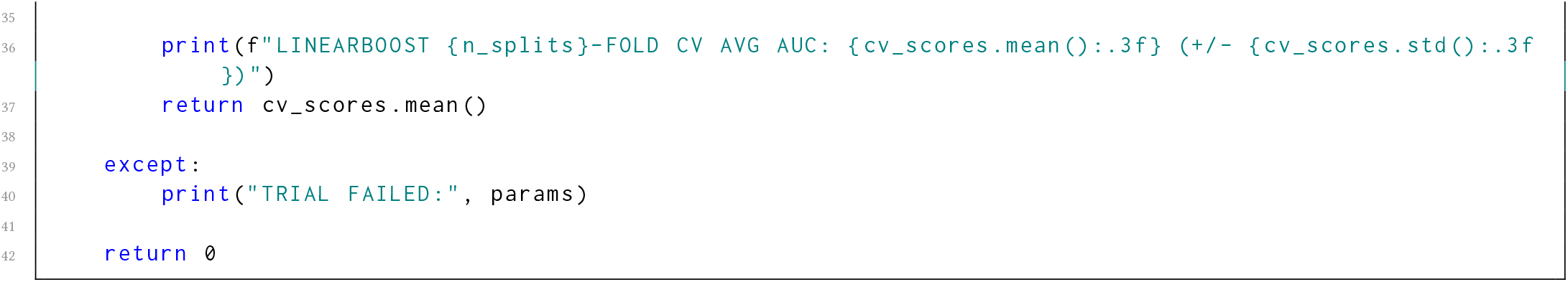
Optuna Hyperparameters for LinearBoost.

**Listing 3.**
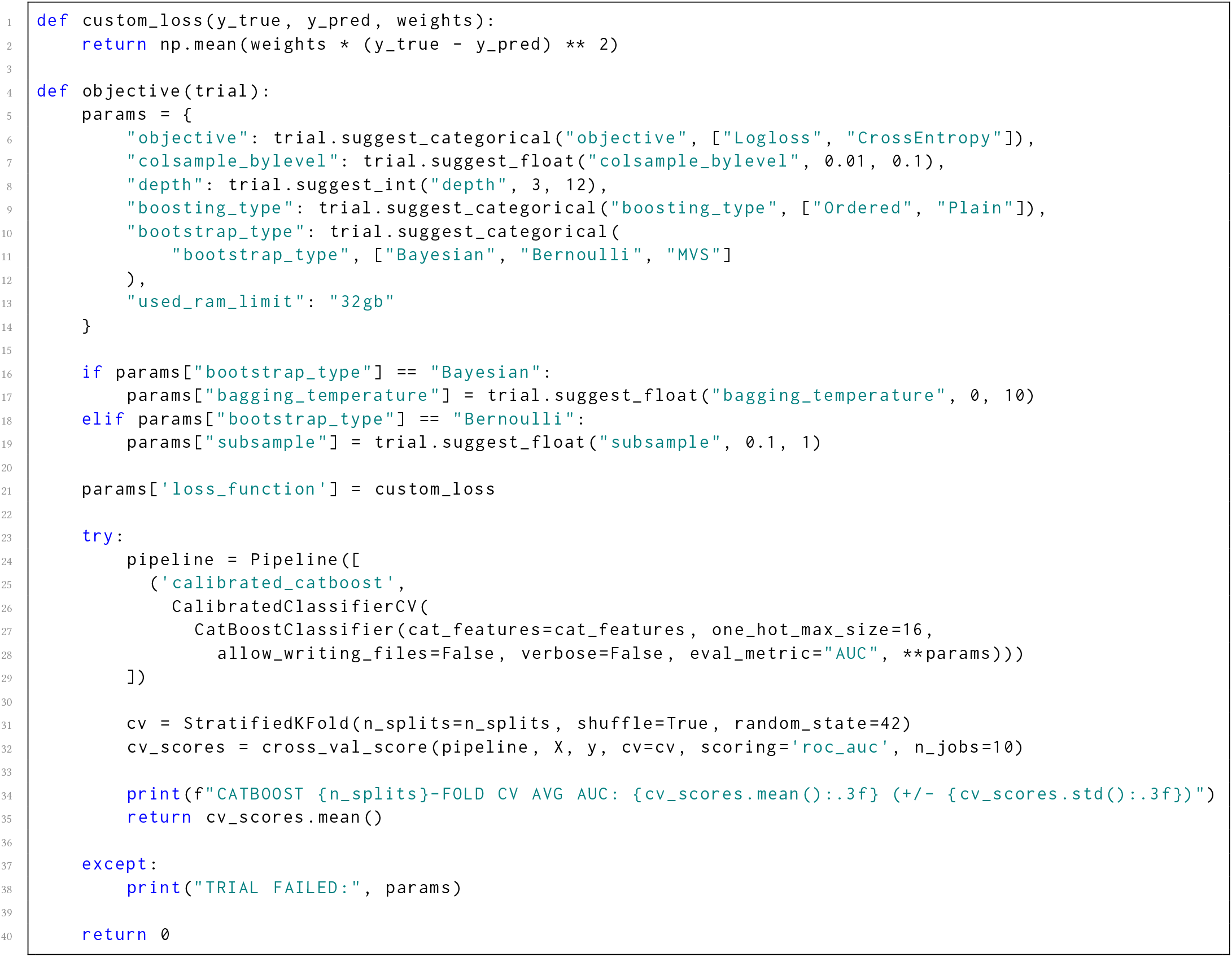
Optuna Hyperparameters for CatBoost.

**Listing 4.**
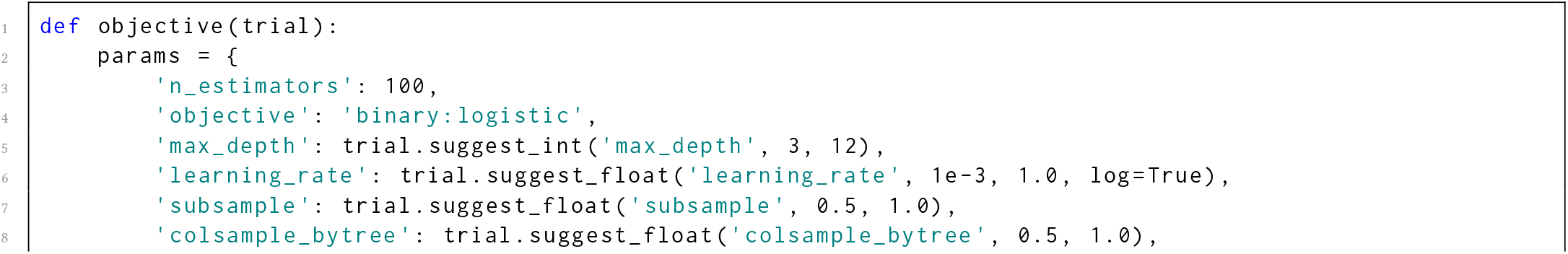
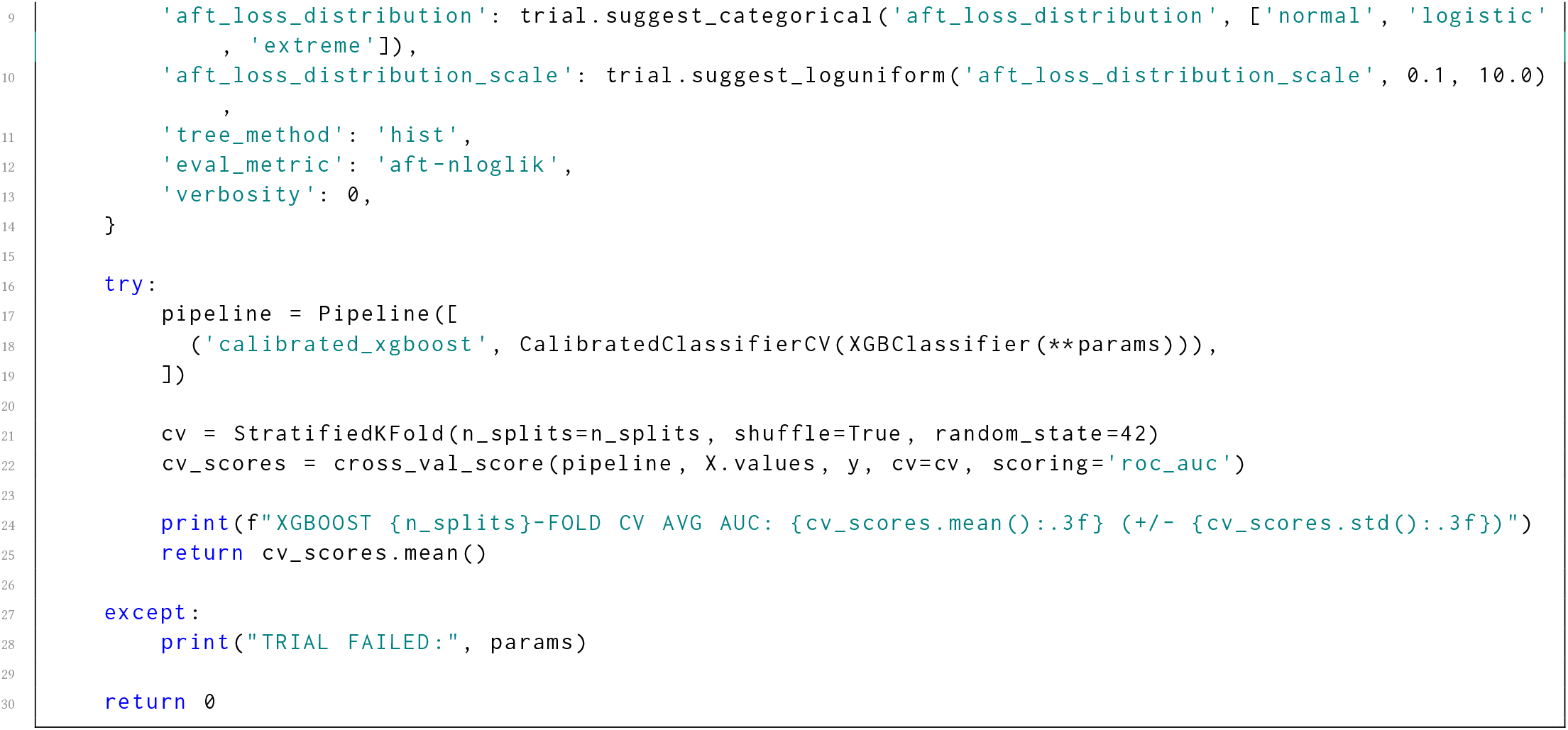
Optuna Hyperparameters for XGBoost.

**Listing 5.**
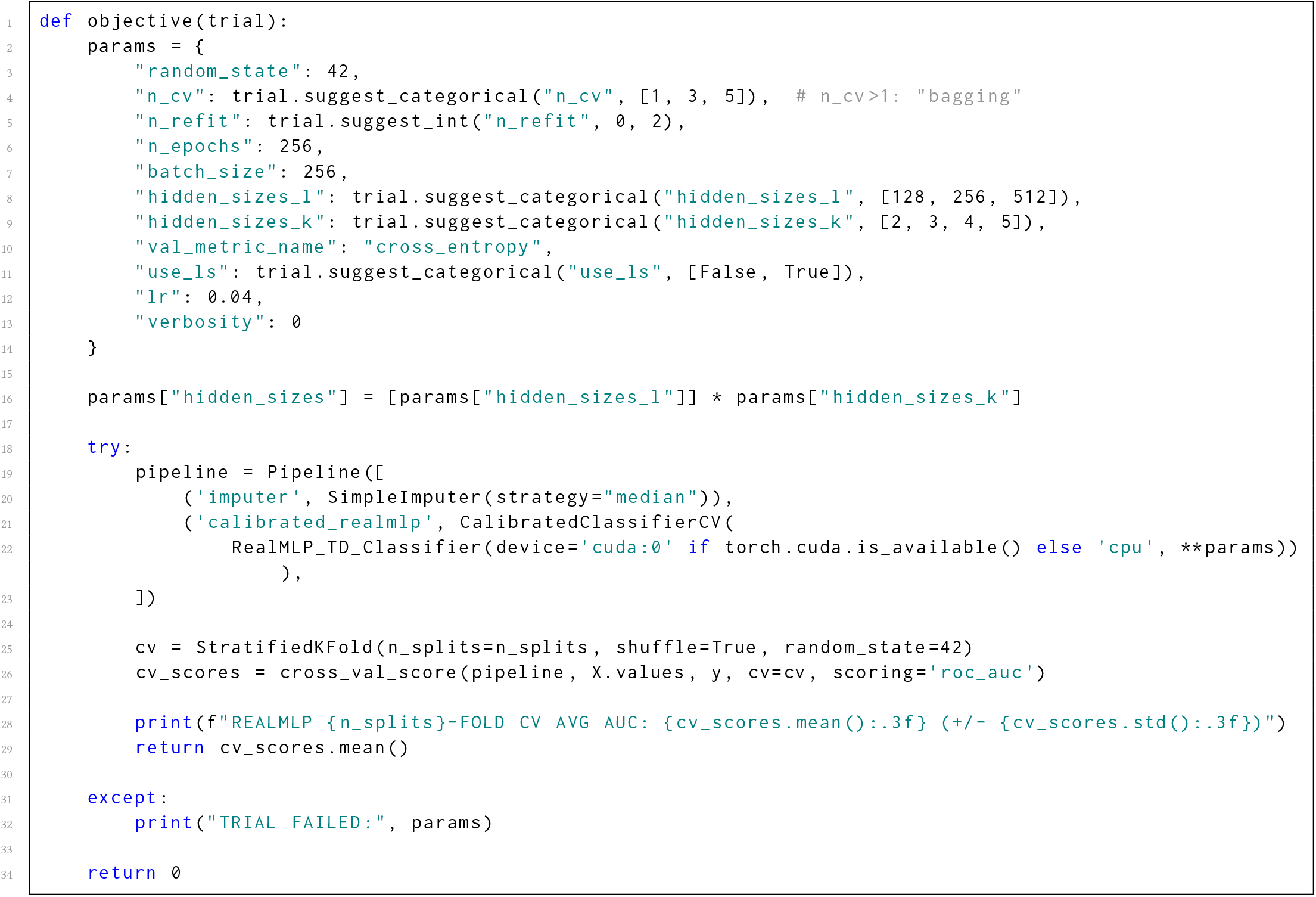
Optuna Hyperparameters for RealMLP.

## Footnotes

1 https://huggingface.co/blog/leaderboard-medicalllm

2 https://cordis.europa.eu/docs/results/305/305739/final1-final-report-riskycad-brief2.pdf

3 https://github.com/dholzmueller/pytabkit

4 https://github.com/LinearBoost/linearboost-classifier

5 https://github.com/catboost/catboost

6 https://github.com/dmlc/xgboost

7 https://github.com/scikit-learn/scikit-learn/tree/main/sklearn/svm

8 https://optuna.org

9 https://huggingface.co/google/medgemma-27b-text-it

10 https://huggingface.co/m42-health/Llama3-Med42-70B

11 https://huggingface.co/Henrychur/MMed-Llama3.1-70B

12 https://huggingface.co/johnsnowlabs/JSL-MedLlama-3-8B-v2.0

13 https://huggingface.co/epfl-llm/meditron-70b

14 https://huggingface.co/aaditya/Llama3-OpenBioLLM-70B

15 https://huggingface.co/meta-llama/Llama-3.1-70B

16 https://github.com/vllm-project/vllm

17 https://github.com/deepspeedai/DeepSpeed

## References

[1] DM Anisuzzaman, Jeffrey G Malins, Paul A Friedman, and Zachi I Attia. 2025. Fine-Tuning Large Language Models for Specialized Use Cases. Mayo Clinic Proceedings: Digital Health 3, 1 (2025), 100184.

[2] Jinze Bai, Shuai Bai, Yunfei Chu, Zeyu Cui, Kai Dang, Xiaodong Deng, Yang Fan, Wenbin Ge, Yu Han, Fei Huang, Binyuan Hui, Luo Ji, Mei Li, Junyang Lin, Runji Lin, Dayiheng Liu, Gao Liu, Chengqiang Lu, Keming Lu, Jianxin Ma, Rui Men, Xingzhang Ren, Xuancheng Ren, Chuanqi Tan, Sinan Tan, Jianhong Tu, Peng Wang, Shijie Wang, Wei Wang, Shengguang Wu, Benfeng Xu, Jin Xu, An Yang, Hao Yang, Jian Yang, Shusheng Yang, Yang Yao, Bowen Yu, Hongyi Yuan, Zheng Yuan, Jianwei Zhang, Xingxuan Zhang, Yichang Zhang, Zhenru Zhang, Chang Zhou, Jingren Zhou, Xiaohuan Zhou, and Tianhang Zhu. 2023. Qwen Technical Report. https://arxiv.org/abs/2309.16609

[3] Himanshu Batra, Narinder Singh Punn, Sanjay Kumar Sonbhadra, and Sonali Agarwal. 2021. BERT-Based Sentiment Analysis: A Software Engineering Perspective. Springer International Publishing, 138–148. doi:10.1007/978-3-030-86472-9_13

[4] Parishad BehnamGhader, Vaibhav Adlakha, Marius Mosbach, Dzmitry Bahdanau, Nicolas Chapados, and Siva Reddy. 2024. LLM2Vec: Large Language Models Are Secretly Powerful Text Encoders. https://arxiv.org/abs/2404.05961

[5] Ofir Ben Shoham and Nadav Rappoport. 2024. CPLLM: Clinical prediction with large language models. PLOS Digital Health 3, 12 (2024), e0000680.

[6] Igor Bibi, Daniel Schaffert, Philipp Blanke, Lorenz Illian, Federico Lenzing, Niklas Martin, Jan Leipe, Winfried März, Ksenija Stach, and Victor Olsavszky. 2025. Cardiovascular risk assessment enhanced by automated machine learning in a multi-phase study. Nature Scientific Reports 15, 1 (2025), 36474

[7] Chaochao Chen, Yuwei Zheng, Xuefen Li, Bo Shen, and Xiaojie Bi. 2025. Biomarkers for Early Predicting In-Hospital Mortality in Severe Fever with Thrombocytopenia Syndrome and Differentiating It from Hemorrhagic Fever with Renal Syndrome. Infection and Drug Resistance 18 (2025), 1355–1366.

[8] Tianqi Chen and Carlos Guestrin. 2016. XGBoost: A Scalable Tree Boosting System. In SIGKDD. 785–794.

[9] DeepSeek-AI, Aixin Liu, Bei Feng, Bing Xue, Bingxuan Wang, Bochao Wu, Chengda Lu, Chenggang Zhao, Chengqi Deng, Chenyu Zhang, Chong Ruan, Damai Dai, Daya Guo, Dejian Yang, Deli Chen, Dongjie Ji, Erhang Li, Fangyun Lin, Fucong Dai, Fuli Luo, Guangbo Hao, Guanting Chen, Guowei Li, H. Zhang, Han Bao, Hanwei Xu, Haocheng Wang, Haowei Zhang, Honghui Ding, Huajian Xin, Huazuo Gao, Hui Li, Hui Qu, J. L. Cai, Jian Liang, Jianzhong Guo, Jiaqi Ni, Jiashi Li, Jiawei Wang, Jin Chen, Jingchang Chen, Jingyang Yuan, Junjie Qiu, Junlong Li, Junxiao Song, Kai Dong, Kai Hu, Kaige Gao, Kang Guan, Kexin Huang, Kuai Yu, Lean Wang, Lecong Zhang, Lei Xu, Leyi Xia, Liang Zhao, Litong Wang, Liyue Zhang, Meng Li, Miaojun Wang, Mingchuan Zhang, Minghua Zhang, Minghui Tang, Mingming Li, Ning Tian, Panpan Huang, Peiyi Wang, Peng Zhang, Qiancheng Wang, Qihao Zhu, Qinyu Chen, Qiushi Du, R. J. Chen, R. L. Jin, Ruiqi Ge, Ruisong Zhang, Ruizhe Pan, Runji Wang, Runxin Xu, Ruoyu Zhang, Ruyi Chen, S. S. Li, Shanghao Lu, Shangyan Zhou, Shanhuang Chen, S. Kom Sande et al. Shaoqing Wu, Shengfeng Ye, Shengfeng Ye, Shirong Ma, Shiyu Wang, Shuang Zhou, Shuiping Yu, Shunfeng Zhou, Shuting Pan, T. Wang, Tao Yun, Tian Pei, Tianyu Sun, W. L. Xiao, Wangding Zeng, Wanjia Zhao, Wei An, Wen Liu, Wenfeng Liang, Wenjun Gao, Wenqin Yu, Wentao Zhang, X. Q. Li, Xiangyue Jin, Xianzu Wang, Xiao Bi, Xiaodong Liu, Xiaohan Wang, Xiaojin Shen, Xiaokang Chen, Xiaokang Zhang, Xiaosha Chen, Xiaotao Nie, Xiaowen Sun, Xiaoxiang Wang, Xin Cheng, Xin Liu, Xin Xie, Xingchao Liu, Xingkai Yu, Xinnan Song, Xinxia Shan, Xinyi Zhou, Xinyu Yang, Xinyuan Li, Xuecheng Su, Xuheng Lin, Y. K. Li, Y. Q. Wang, Y. X. Wei, Y. X. Zhu, Yang Zhang, Yanhong Xu, Yanhong Xu, Yanping Huang, Yao Li, Yao Zhao, Yaofeng Sun, Yaohui Li, Yaohui Wang, Yi Yu, Yi Zheng, Yichao Zhang, Yifan Shi, Yiliang Xiong, Ying He, Ying Tang, Yishi Piao, Yisong Wang, Yixuan Tan, Yiyang Ma, Yiyuan Liu, Yongqiang Guo, Yu Wu, Yuan Ou, Yuchen Zhu, Yuduan Wang, Yue Gong, Yuheng Zou, Yujia He, Yukun Zha, Yunfan Xiong, Yunxian Ma, Yuting Yan, Yuxiang Luo, Yuxiang You, Yuxuan Liu, Yuyang Zhou, Z. F. Wu, Z. Z. Ren, Zehui Ren, Zhangli Sha, Zhe Fu, Zhean Xu, Zhen Huang, Zhen Zhang, Zhenda Xie, Zhengyan Zhang, Zhewen Hao, Zhibin Gou, Zhicheng Ma, Zhigang Yan, Zhihong Shao, Zhipeng Xu, Zhiyu Wu, Zhongyu Zhang, Zhuoshu Li, Zihui Gu, Zijia Zhu, Zijun Liu, Zilin Li, Ziwei Xie, Ziyang Song, Ziyi Gao, and Zizheng Pan. 2025. DeepSeek-V3 Technical Report. https://arxiv.org/abs/2412.19437

[10] Jacob Devlin, Ming-Wei Chang, Kenton Lee, and Kristina Toutanova. 2019. BERT: Pre-training of Deep Bidirectional Transformers for Language Understanding. In NAACL-HLT. 4171–4186.

[11] Mariachiara Di Cesare, Pablo Perel, Sean Taylor, Chodziwadziwa Kabudula, Honor Bixby, Thomas A. Gaziano, Diana Vaca McGhie, Jeremiah Mwangi, Borjana Pervan, Jagat Narula, Daniel Pineiro, and Fausto J. Pinto. 2024. The Heart of the World. Global Heart 19, 1 (2024), 11.

[12] Nick Erickson, Lennart Purucker, Andrej Tschalzev, David Holzmüller, Prateek Mutalik Desai, David Salinas, and Frank Hutter. 2025. TabArena: A Living Benchmark for Machine Learning on Tabular Data. https://arxiv.org/abs/2506.16791

[13] Rean Fernandes, André Biedenkapp, Frank Hutter, and Noor Awad. 2025. A Llama walks into the ‘Bar’: Efficient Supervised Fine-Tuning for Legal Reasoning in the Multi-state Bar Exam. https://arxiv.org/abs/2504.04945

[14] Chuan Guo, Geoff Pleiss, Yu Sun, and Kilian Q. Weinberger. 2017. On Calibration of Modern Neural Networks. In PMLR. 1321–1330.

[15] Changho Han, Dong Won Kim, Songsoo Kim, Seng Chan You, Jin Young Park, SungA Bae, and Dukyong Yoon. 2024. Evaluation of GPT-4 for 10-year cardiovascular risk prediction: Insights from the UK Biobank and KoGES data. iScience 27, 2 (2024), 109022.

[16] Aron Henriksson, Yash Pawar, Pontus Hedberg, and Pontus Nauclér. 2023. Multimodal fine-tuning of clinical language models for predicting COVID-19 outcomes. Artificial Intelligence in Medicine 146 (2023), 102695.

[17] Guillem Hernández Guillamet, Ariadna Ning Morancho Pallaruelo, Laura Miró Mezquita, Ramón Miralles, Miquel Àngel Mas, María José Ulldemolins Papaseit, Oriol Estrada Cuxart, and Francesc López Seguí. 2023. Machine Learning Model for Predicting Mortality Risk in Patients With Complex Chronic Conditions: Retrospective Analysis. Online Journal of Public Health Informatics 15 (2023).

[18] Noah Hollmann, Samuel Müller, Lennart Purucker, Arjun Krishnakumar, Max Körfer, Shi Bin Hoo, Robin Tibor Schirrmeister, and Frank Hutter. 2025. Accurate predictions on small data with a tabular foundation model. Nature 637, 8045 (2025), 319–326.

[19] David Holzmüller, Léo Grinsztajn, and Ingo Steinwart. 2024. Better by default: Strong pre-tuned MLPs and boosted trees on tabular data. In NeurIPS. 26577–26658.

[20] Edward J. Hu, Yelong Shen, Phillip Wallis, Zeyuan Allen-Zhu, Yuanzhi Li, Shean Wang, Lu Wang, and Weizhu Chen. 2021. LoRA: Low-Rank Adaptation of Large Language Models. https://arxiv.org/abs/2106.09685

[21] Siva Rajesh Kasa, Karan Gupta, Sumegh Roychowdhury, Ashutosh Kumar, Yaswanth Biruduraju, Santhosh Kumar Kasa, Pattisapu Nikhil Priyatam, Arindam Bhattacharya, Shailendra Agarwal, and Vijay Huddar. 2025. Generative or Discriminative? Revisiting Text Classification in the Era of Transformers. In EMNLP. 9604–9626.

[22] Guolin Ke, Qi Meng, Thomas Finley, Taifeng Wang, Wei Chen, Weidong Ma, Qiwei Ye, and Tie-Yan Liu. 2017. LightGBM: A Highly Efficient Gradient Boosting Decision Tree. In NeurIPS, Isabelle Guyon, Ulrike von Luxburg, Samy Bengio, Hanna M. Wallach, Rob Fergus, S. V. N. Vishwanathan, and Roman Garnett (Eds.). 3146–3154.

[23] Joseph Lee, Tianqi Shang, Jae Young Baik, Duy Duong-Tran, Shu Yang, Lingyao Li, and Li Shen. 2025. From Promising Capability to Pervasive Bias: Assessing Large Language Models for Emergency Department Triage. https://arxiv.org/abs/2504.16273

[24] Dubai Li, Nan Jiang, Kangping Huang, Ruiqi Tu, Shuyu Ouyang, Huayu Yu, Lin Qiao, Chen Yu, Tianshu Zhou, Danyang Tong, Qian Wang, Mengtao Li, Xiaofeng Zeng, Yu Tian, Xinping Tian, and Jingsong Li. 2025. From Questions to Clinical Recommendations: Large Language Models Driving Evidence-Based Clinical Decision Making. https://arxiv.org/abs/2505.10282

[25] Lingyao Li, Jiayan Zhou, Zhenxiang Gao, Wenyue Hua, Lizhou Fan, Huizi Yu, Loni Hagen, Yongfeng Zhang, Themistocles L. Assimes, Libby Hemphill, and Siyuan Ma. 2024. A scoping review of using Large Language Models (LLMs) to investigate Electronic Health Records (EHRs). https://arxiv.org/abs/2405.03066

[26] Daniel Lindholm, Johan Lindbäck, Paul W. Armstrong, Andrzej Budaj, Christopher P. Cannon, Christopher B. Granger, Emil Hagström, Claes Held, Wolfgang Koenig, Ollie Östlund, Ralph A.H. Stewart, Joseph Soffer, Harvey D. White, Robbert J. De Winter, Philippe Gabriel Steg, Agneta Siegbahn, Marcus E. Kleber, Alexander Dressel, Tanja B. Grammer, Winfried März, and Lars Wallentin. 2017. Biomarker-Based Risk Model to Predict Cardiovascular Mortality in Patients With Stable Coronary Disease. Journal of the American College of Cardiology 70, 7 (2017), 813–826.

[27] Yang Liu. 2019. Fine-tune BERT for Extractive Summarization. https://arxiv.org/abs/1903.10318

[28] Subhankar Maity and Manob Jyoti Saikia. 2025. Large Language Models in Healthcare and Medical Applications: A Review. Bioengineering 12, 6 (2025), 631.

[29] Nikita Makarov, Maria Bordukova, Papichaya Quengdaeng, Daniel Garger, Raul Rodriguez-Esteban, Fabian Schmich, and Michael P. Menden. 2025. Large language models forecast patient health trajectories enabling digital twins. Digital Medicine 8, 1 (2025), 588.

[30] Moman A Mohammad, Kevin KW Olesen, Sasha Koul, Chris P Gale, Rebecca Rylance, Tomas Jernberg, Tomasz Baron, Jonas Spaak, Stefan James, Bertil Lindahl, and others. 2022. Development and validation of an artificial neural network algorithm to predict mortality and admission to hospital for heart failure after myocardial infarction: a nationwide population-based study. The Lancet Digital Health 4, 1 (2022), e37–e45.

[31] Parvati Naliyatthaliyazchayil, Raajitha Muthyala, Judy Wawira Gichoya, and Saptarshi Purkayastha. 2025. Evaluating the Reasoning Capabilities of Large Language Models for Medical Coding and Hospital Readmission Risk Stratification: Zero-Shot Prompting Approach. Journal of Medical Internet Research 27 (2025), e74142.

[32] Charles O’Neill, Tirthankar Ghosal, Roberta Răileanu, Mike Walmsley, Thang Bui, Kevin Schawinski, and Ioana Ciucă. 2025. Sparks of Science: Hypothesis Generation Using Structured Paper Data. https://arxiv.org/abs/2504.12976

[33] Yash Pawar, Aron Henriksson, Pontus Hedberg, and Pontus Naucler. 2022. Leveraging clinical BERT in multimodal mortality prediction models for COVID-19. In CBMS. 199–204.

[34] John Platt. 1999. Probabilistic outputs for support vector machines and comparisons to regularized likelihood methods. Adv. Large Margin Classifiers 10, 3 (1999), 61–74.

[35] Liudmila Prokhorenkova, Gleb Gusev, Aleksandr Vorobev, Anna Veronika Dorogush, and Andrey Gulin. 2018. CatBoost: unbiased boosting with categorical features. In NIPS. 6639–6649.

[36] Alireza Salemi and Hamed Zamani. 2025. LaMP-QA: A Benchmark for Personalized Long-form Question Answering. https://arxiv.org/abs/2506.00137

[37] SCORE2 Working Group. 2021. SCORE2 risk prediction algorithms: new models to estimate 10-year risk of cardiovascular disease in Europe. European Heart Journal 42, 25 (2021), 2439–2454.

[38] Boqun Shi et al. 2025. Large language models and artificial neural networks for assessing 1-year mortality in patients with myocardial infarction: Analysis from the Medical Information Mart for Intensive Care IV (MIMIC-IV) database. Journal of Medical Internet Research 27 (2025).

[39] SMART Study Group. 2013. Development and validation of a prediction rule for recurrent vascular events based on a cohort study of patients with arterial disease: the SMART risk score. Heart (British Cardiac Society) 99, 12 (2013), 866–872.

[40] Hugo Touvron, Thibaut Lavril, Gautier Izacard, Xavier Martinet, Marie-Anne Lachaux, Timothée Lacroix, Baptiste Rozière, Naman Goyal, Eric Hambro, Faisal Azhar, Aurelien Rodriguez, Armand Joulin, Edouard Grave, and Guillaume Lample. 2023. LLaMA: Open and Efficient Foundation Language Models. https://arxiv.org/abs/2302.13971

[41] Konstantina Tsarapatsani, Antonis I. Sakellarios, Vasileios C. Pezoulas, Vasilis D. Tsakanikas, George K. Matsopoulos, Winfried März, Marcus E. Kleber, and Dimitrios I. Fotiadis. 2022. Machine Learning Models to Predict Myocardial Infarction Within 10-Years Follow-up of Cardiovascular Disease Progression. In EMBS. 1–4.

[42] Konstantina Tsarapatsani, Antonis I. Sakellarios, Vasilis D. Tsakanikas, Marcus E. Kleber, Winfried März, and Dimitrios I. Fotiadis. 2023. Machine Learning Models Predict Fatal Myocardial Infarction Within 10-Years Follow-Up Utilizing Explainable AI. In BIBE. 320–324.

[43] Ashish Vaswani, Noam Shazeer, Niki Parmar, Jakob Uszkoreit, Llion Jones, Aidan N. Gomez, Lukasz Kaiser, and Illia Polosukhin. 2017. Attention is All you Need. In NeurIPS. 5998–6008.

[44] Xinyu Wang, Haonan Wang, and Rui Fu. 2025. Boosting Cardiovascular Disease Prediction Accuracy: A Hybrid AI Strategy Integrating LLM-Generated Risk Scores. In HIDA. 179–185.

[45] Bernhard R Winkelmann, Winfried März, Bernhard O Boehm, Rainer Zotz, Jörg Hager, Peter Hellstern, and Jochen Senges. 2001. Rationale and design of the LURIC study-a resource for functional genomics, pharmacogenomics and long-term prognosis of cardiovascular disease. Pharmacogenomics 2, 1 (2001), S1–73.

[46] Haolun Wu, Ye Yuan, Liana Mikaelyan, Alexander Meulemans, Xue Liu, James Hensman, and Bhaskar Mitra. 2024. Learning to Extract Structured Entities Using Language Models. https://arxiv.org/abs/2402.04437

[47] Zhen Wu, Abdullahi Mohamud Hilowle, Ying Zhou, Changlin Zhao, and Shuo Yang. 2025. Delving into biomarkers and predictive modeling for CVD mortality: a 20-year cohort study. Nature Scientific Reports 15, 1 (2025), 4134.

[48] An Yang, Anfeng Li, Baosong Yang, Beichen Zhang, Binyuan Hui, Bo Zheng, Bowen Yu, Chang Gao, Chengen Huang, Chenxu Lv, Chujie Zheng, Dayiheng Liu, Fan Zhou, Fei Huang, Feng Hu, Hao Ge, Haoran Wei, Huan Lin, Jialong Tang, Jian Yang, Jianhong Tu, Jianwei Zhang, Jianxin Yang, Jiaxi Yang, Jing Zhou, Jingren Zhou, Junyang Lin, Kai Dang, Keqin Bao, Kexin Yang, L. Yu, Lianghao Deng, Mei Li, Mingfeng Xue, Mingze Li, Pei Zhang, Peng Wang, Qin Zhu, Rui Men, Ruize Gao, Shixuan Liu, Shuang Luo, Tianhao Li, Tianyi Tang, Wenbiao Yin, Xingzhang Ren, Xinyu Wang, Xinyu Zhang, Xuancheng Ren, Yang Fan, Yang Su, Yichang Zhang, Yinger Zhang, Yu Wan, Yuqiong Liu, Zekun Wang, Zeyu Cui, Zhenru Zhang, Zhipeng Zhou, and Zihan Qiu. 2025. Qwen3 Technical Report. https://arxiv.org/abs/2505.09388

[49] Tazveen Zanab et al. 2025. Advancing Cardiovascular Disease Prediction Using LLMs. In ICMLAS. 581–588.

[50] Shuang Zhou et al. 2025. Large language models for disease diagnosis: a scoping review. Artificial Intelligence 1, 1 (2025), 9.

[51] Jinhua Zhu, Yingce Xia, Lijun Wu, D. He, Tao Qin, Wengang Zhou, Houqiang Li, and Tie-Yan Liu. 2020. Incorporating BERT into Neural Machine Translation. https://arxiv.org/abs/2002.06823

